# INTEGRATING GENOMIC AND FUNCTIONAL TESTING TO IMPROVE CFTR MODULATOR RESPONSE PREDICTION IN CHILDREN WITH CYSTIC FIBROSIS

**DOI:** 10.1101/2025.07.09.25331152

**Authors:** Laura K. Fawcett, Zahra A. Chew, Elena K. Schneider-Futschik, Katelin M. Allan, Hardip R. Patel, Adam Jaffe, Shafagh A. Waters

## Abstract

**Background:** CFTR modulators have transformed cystic fibrosis (CF) treatment, but individual responses vary even among patients with identical CFTR genotypes. This underscores the need for predictive biomarkers to optimize therapeutic selection.

**Methods:** We evaluated 24 paediatric patients homozygous for F508del-CFTR, assessing lung function (FEV1pp) and sweat chloride (SC) before and after CFTR modulator therapy. Whole-gene sequencing was utilised to identify CFTR and pharmacogene variants. Patient-derived human nasal epithelial cells (HNECs) were expanded and differentiated at the air-liquid interface to assess CFTR function via ion transport (ΔIsc).

**Results:** Clinical responses varied widely. Twelve participants changed modulators during the study. Sequencing identified 231 additional CFTR variants and pharmacogene polymorphisms, but none correlated with response variability. However, a significant linear relationship emerged between ΔIsc and FEV1pp improvement in patients with baseline FEV1pp <90 (R² = 0.651, p = 0.001) and SC reduction (R² = 0.535, p = 0.004). Receiver operating characteristic (ROC) analysis demonstrated high predictive accuracy for SC reduction (AUC = 0.88) and combined FEV1pp/SC response in patients with baseline FEV1pp <90 (AUC = 1.00). Exploratory analysis confirmed that ΔIsc predicts FEV1pp changes, modulated by baseline lung function and CFTR modulator type.

**Conclusion:** Patient-derived differentiated HNEC cultures serve as a robust predictive tool for CFTR modulator response in paediatric CF patients. Their integration into clinical practice can enhance personalised treatment strategies, minimising ineffective therapy use and improving CF patient outcomes with precision medicine.

**What is already known on this topic:**

- CFTR modulators significantly improve clinical outcomes in people with cystic fibrosis (CF), yet individual responses vary, even among those with identical CFTR genotypes.
- Patient-derived human nasal epithelial cell (HNEC) models have emerged as promising tools to predict CFTR modulator response. However, existing studies have primarily focused on adult and adolescent populations, leaving a gap in personalised treatment strategies for younger children with CF.
- The relationship between CFTR sequence variations, pharmacogene heterogeneity, and modulator response in paediatric patients has not been extensively explored.

**What this study adds:**

- This study demonstrates a strong correlation between in vitro CFTR function (ΔIsc) and clinical improvements in FEV1pp and sweat chloride in children and adolescents with CF.
- Whole-gene sequencing identified **231 additional CFTR variants**, yet none were associated with CFTR modulator response, suggesting that genotype alone does not fully explain treatment variability.
- While some trends between pharmacogene activity and treatment response were observed, no strong evidence supports pharmacogene profiling as a standalone predictor of CFTR modulator efficacy in paediatric patients.
- **Differentiated-HNEC cultures consistently predicted clinical response across multiple CFTR modulator regimens**, reinforcing their value for preclinical drug screening.

**How this study might affect research, practice, or policy:**

- Our findings support the integration of **differentiated-HNEC models into clinical practice** to personalise CFTR modulator selection, reducing ineffective treatments and improving patient outcomes.
- The study underscores the **need for additional clinical endpoints** beyond FEV1pp to assess respiratory function in individuals with preserved lung function (FEV1pp > 90%).

## INTRODUCTION

Cystic Fibrosis (CF) is an autosomal recessive disorder caused by pathological variants in the CF transmembrane conductance regulator (CFTR) gene, that encodes a chloride and bicarbonate channel ^1^. The most prevalent CF-causing variant, F508del, results in CFTR protein misfolding, premature degradation, and impaired gating function ^2,3^. The development of CFTR modulators, such as potentiators like ivacaftor (IVA; I) and correctors such as lumacaftor (LUM), tezacaftor (TEZ; T), and elexacaftor (E) has revolutionised treatment for people with CF (PwCF). These modulators, when used as dual or triple therapy, target specific CFTR variants like F508del, to address the multifactorial dysfunction caused by this variant ^4,5^.

Implementation of CFTR modulators to the clinic faces challenges due to the considerable heterogeneity in clinical responses between PwCF ^6–9^. Current clinical tools to assess response to CFTR modulator therapy include measurement of forced expiratory volume in one second (FEV1) and sweat chloride (SC) levels. FEV1 has been the primary endpoint in most phase III trials, however its limitations, such as reduced sensitivity to early lung disease changes, has led to exclusion of individuals with FEV1 values above 90 percent predicted (pp) ^9–11^. SC, the diagnostic gold standard, is now also used as a biomarker for CFTR modulator efficacy ^12^. Changes in clinical endpoints are continuous, however an increase in FEV1pp by more than 5 percentage points and reduction in SC by more than 20mmol/L have typically been accepted to indicate a significant positive therapeutic response to treatment ^12–15^.

Accurately predicting the clinical response is important because incorrect or suboptimal treatments may lead to increased healthcare costs and reduced quality of life due to adverse side effects ^1^. When variability in CFTR modulator efficacy between PwCF is not due to their adherence to treatment, it may be explained by drug-drug interactions with concomitant CF therapies ^16,17^. Alternatively, genetic modifiers ^18^ such as polymorphisms in drug metabolising enzymes, pharmacogenes, can result in poor metabolisers or ultra-rapid metabolisers of CFTR modulators ^17,19^. Complex CFTR alleles, additional variants present with F508del, have also been shown to impact clinical response to CFTR modulators ^20,21^. Standard clinical CFTR genotyping may overlook these additional variants, highlighting the importance of comprehensive genetic profiling to accurately identify responders.

To address the challenge of predicting individual responses to CFTR modulators, preclinical methodologies using patient-derived primary cell models have emerged as promising tools, which are also acceptable to patients ^22–24^. These models, derived from the airway (nasal, bronchial) and intestinal epithelial cells, enable characterisation of the molecular and functional defects caused by CFTR variants in a biologically relevant system, as well as assessment of a therapy’s efficacy in modulating CFTR protein activity ^25,26^. Differentiation of human airway cells into pseudostratified epithelium at air-liquid interface (ALI) facilitates direct measurement of ion transport, enabling accurate evaluation of CFTR function and drug efficacy ^27–31^. Differentiated-human nasal epithelial cells (HNECs), validated for CF modulator drug testing, offer non-invasive accessibility compared to bronchial cells ^27^. The gold standard for evaluating CFTR function involves electrophysiological ion transport assessments, specifically measuring short-circuit-current (Isc) in the Ussing chamber to detect changes in cAMP-mediated chloride (Cl^-^) transport induced by CFTR modulators ^32,33^.

Several studies have demonstrated a strong positive correlation between ΔIsc measurements in paired differentiated-HNEC cultures and the clinical response to CFTR modulators, in both common and rare CFTR variants (**Table S1**) ^31,34–36^. Pranke (2017, 2019) and Debley (2020) found a strong correlation between ΔIsc in differentiated-HNEC cultures and changes in FEV1pp, and SC in PwCF with F508del-CFTR and gating variants treated with LUM/IVA and IVA, respectively ^31,35,36^. Dreano (2023) reported similar correlations in a diverse cohort of non F508del-CFTR PwCF ^34^. Additional studies have provided individual results for those with rare CF-causing CFTR variants, demonstrating the broader applicability of these findings ^37–39^.

Although insightful, these studies are limited by a small number of participants, primarily adults and adolescents. Early CFTR modulator treatment can profoundly affect disease progression, making young PwCF prime candidates for guiding modulator selection due to the potential for long-term benefits. In this study, the relationships between *in vivo* clinical response (FEV1pp and SC), genetics (CFTR and pharmacogenes) and *in vitro* functional response (ΔIsc in differentiated-HNEC) were investigated (**Figure 1**).

**Figure 1:**
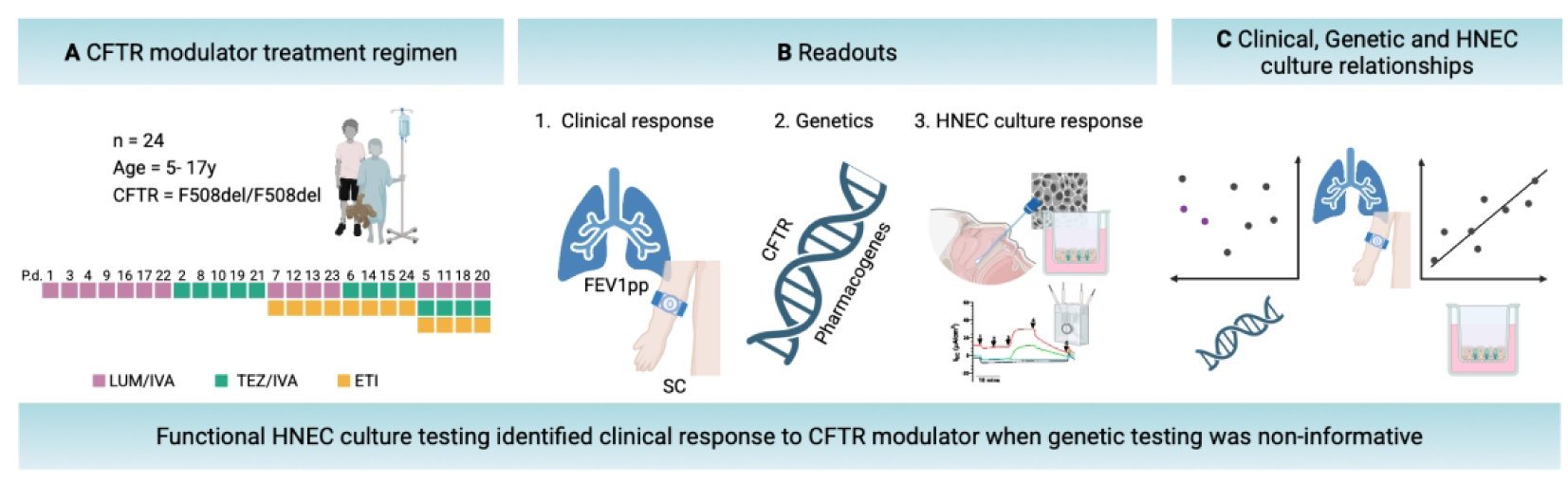
Overview of study design and analytical framework A) CFTR modulator treatment regimen. Each box on the first row represents one of the 24 study participants (F508del/F508del CFTR, Aged 5-17 years) P.d; Participant de-identifier. Subsequent rows illustrate changes in treatment regimen, denoted by the switch in colour within columns. The colour indicates the specific CFTR modulators administered, including LUM/IVA (lumacaftor/ivacaftor), TEZ/IVA (tezacaftor/ivacaftor), or ETI (elexacaftor/tezacaftor/ivacaftor). **B) Readouts.** *Clinical response (in vivo)*: measured via lung function (FEV1pp) and sweat chloride (SC) pre- and post-treatment. *Genetics:* Comprehensive CFTR and pharmacogene sequencing. *Human Nasal Epithelial Cell (HNEC) culture response (in vitro):* assessed using electrophysiological measurements of CFTR function in differentiated-HNEC cultures derived from each participant. C) Clinical, Genetic and HNEC culture Relationships. Clinical outcomes, including FEV1 percent predicted and sweat chloride levels, are compared with genetic sequencing data and to differentiated-HNEC culture responses to CFTR modulators to evaluate the predictive value of epithelial cell models. FEV1pp: Forced Expiratory Volume in 1 second, percent predicted. Created in BioRender. Waters, S. (2025) https://BioRender.com/v19c483 457×139mm (118 × 118 DPI)

## MATERIAL AND METHODS (Supplementary file #2)

Detailed material and methods are available as supplementary materials.

## RESULTS

### Baseline characteristics and modulator treatment regimen

In this study, 24 PwCF aged between 5 and 17 years, all homozygous for the F508del-CFTR variant met the inclusion criteria (**Table 1, Table S2**). Prior to treatment with modulator therapy (baseline), every participant exhibited an FEV1pp above 40, with half of the group having an FEV1pp above 90 (**Table 1, Table S2**). Each participant received an age-appropriate CFTR modulator as part of their comprehensive CF management with 15 participants receiving LUM/IVA and 9 participants receiving TEZ/IVA. Twelve participants underwent changes in their CFTR modulator regimen during the study period. Four participants transitioned directly from LUM/IVA to ETI, another four from TEZ/IVA to ETI and the remaining four transitioned from LUM/IVA to TEZ/IVA before subsequently transitioning to ETI (**Figure 1A**). Measurement of CFTR modulator levels in participants’ blood samples confirmed adherence to the prescribed regimen, with median LUM detected 48.35 µg/mL and median TEZ detected 47.88 µg/mL (**Figure S1**).

### Heterogenous *in vivo* clinical response (FEV1pp and SC)

The clinical response of all 24 participants to their first CFTR modulator was evaluated at least 6 weeks after starting treatment (*Material and Methods*). The mean increase in FEV1pp after treatment with LUM/IVA or TEZ/IVA was 3.56 percentage points (95% CI; 0.40 - 6.72, **Figure 2A**). The mean decrease in SC after treatment was 21.8 mmol/L (95% CI; 15.81 - 27.76, **Figure 2B**). Consistent with previous studies ^7–9^, there was marked heterogeneity in both FEV1pp and SC response to modulators between individual participants (**Figure 2**).

**Figure 2:**
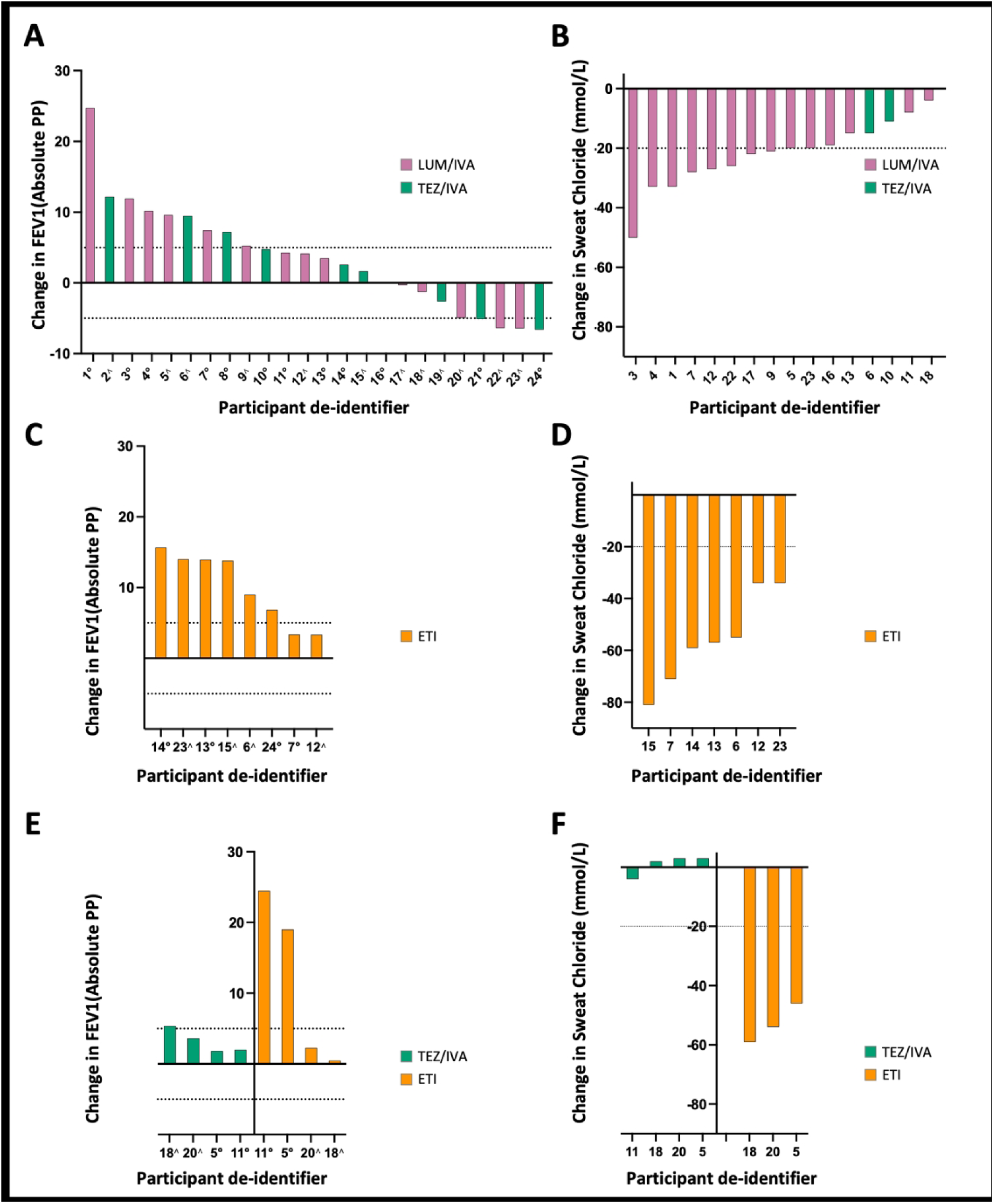
Heterogenous in vivo clinical response to CFTR modulators. Waterfall plots illustrate the absolute change in participant outcomes after treatment with CFTR modulators. **A) Change in FEV1pp.** Absolute change in FEV1pp following treatment with lumacaftor/ivacaftor (LUM/IVA) or tezacaftor/ivacaftor (TEZ/IVA). Each bar represents an individual participant, and dotted lines indicate significant changes from baseline. **B) Change in SC**: Absolute change in SC levels after treatment with LUM/IVA or TEZ/IVA. Each bar represents a participant, with significant changes indicated by dotted lines. **C) Subsequent Change in FEV1**: Absolute change in FEV1pp after switching directly to elexacaftor/tezacaftor/ivacaftor (ETI). Change is calculated as the difference between the subsequent baseline following treatment with a modulator, taken immediately prior to the new modulator, and post treatment values for the new regimen. **D) Subsequent Change in SC:** Absolute change in SC levels after switching directly to ETI, calculated as the difference between baseline (no treatment) and post-treatment values for the new regimen. **E) Subsequent Change in FEV1 in participants who transitioned from LUM/IVA to TEZ/IVA before switching to ETI**: Absolute change in FEV1pp after switching to a subsequent CFTR modulator TEZ/IVA or elexacaftor/tezacaftor/ivacaftor (ETI). Change is calculated as the difference between the subsequent baseline following treatment with a modulator, taken immediately prior to the new modulator, and post treatment values for the new regimen **F) Subsequent Change in SC in participants who transitioned from LUM/IVA to TEZ/IVA before switching to ETI**: Absolute change in SC levels after switching to TEZ/IVA or ETI, calculated as the difference between baseline (no treatment) and post-treatment values for the new regimen. Participants are identified consistently across all four graphs using a participant de-identifier. Nine participants had incomplete SC data (**Table S2**). Symbols denote participants with a baseline FEV1pp below 90 (∘) or above 90 (∧). FEV1pp: Forced Expiratory Volume in 1 second, percent predicted. SC: Sweat Chloride. 182×220mm (300 × 300 DPI)

Next, we evaluated the clinical response of the eight participants who switched their modulator treatment directly to ETI. After treatment with ETI, FEV1pp increased by a mean of 10.00 percentage points (95% CI 5.79 – 14.21, **Figure 2C**), from their subsequent baseline following prior modulator treatment, although the response remained heterogenous between participants (**Figure S2**). All participants had a significant (> 20mmol/L) decrease in SC compared to their original baseline levels, although the variability in the magnitude of response was large (Mean 55.86 mmol/L, 95% CI; 39.71 – 72.00, **Figure 2D**). Two participants had minimal additional improvement in SC with ETI compared to their first modulator (**Figure S2**).

We also evaluated the clinical response of the four participants who transitioned from LUM/IVA to TEZ/IVA before switching to ETI. Only one participant showed a significant increase in FEV1pp with TEZ/IVA treatment (#18: 5.34 percentage points, **Figure 2E, S2**). However, there were no significant (> 20mmol/L) changes in SC levels compared to baseline for any of the four participants (**Figure 2F, S2**). With ETI treatment, two participants had significant increase in FEV1pp (#5: 19.00 percentage points; #11: 24.47 percentage points, **Figure 2E, S2**). The two participants without significant change in FEV1pp had baseline measurements above 90 (#18 and #20). Three participants who had available SC data (#18, #20 and #5) showed a significant (> 20mmol/L) decrease in SC levels, ranging from a 46 to 59 mmol/L decrease compared to baseline (**Figure 2F, S2**).

### Lack of correlation between additional CFTR variants and clinical response (FEV1pp and SC)

Given that additional variants alongside F508del could affect modulator effectiveness, we investigated these variants in our cohort ^40^. Sequencing of the CFTR gene confirmed that all 24 participants were homozygous for F508del, and in addition identified 231 variants across 218 variable sites, from the CFTR promotor to the 3’untranslated region (**Figure 3, S3, Table S3**). Phasing of these variants revealed 44 distinct haplotypes, which reduced to only 3 haplotypes when considering coding sequence only. Twenty participants were homozygous for one haplotype, designated haplotype AA, three carried haplotype AB and one carried haplotype AC (**Table S2**). The CFTR variant L467F, previously reported to make F508del patients non-responsive to modulators was not found in our cohort^20,21,40^. No discernible patterns of clinical response as measured by FEV1pp and SC were observed with the haplotype (**Table S2**).

**Figure 3:**
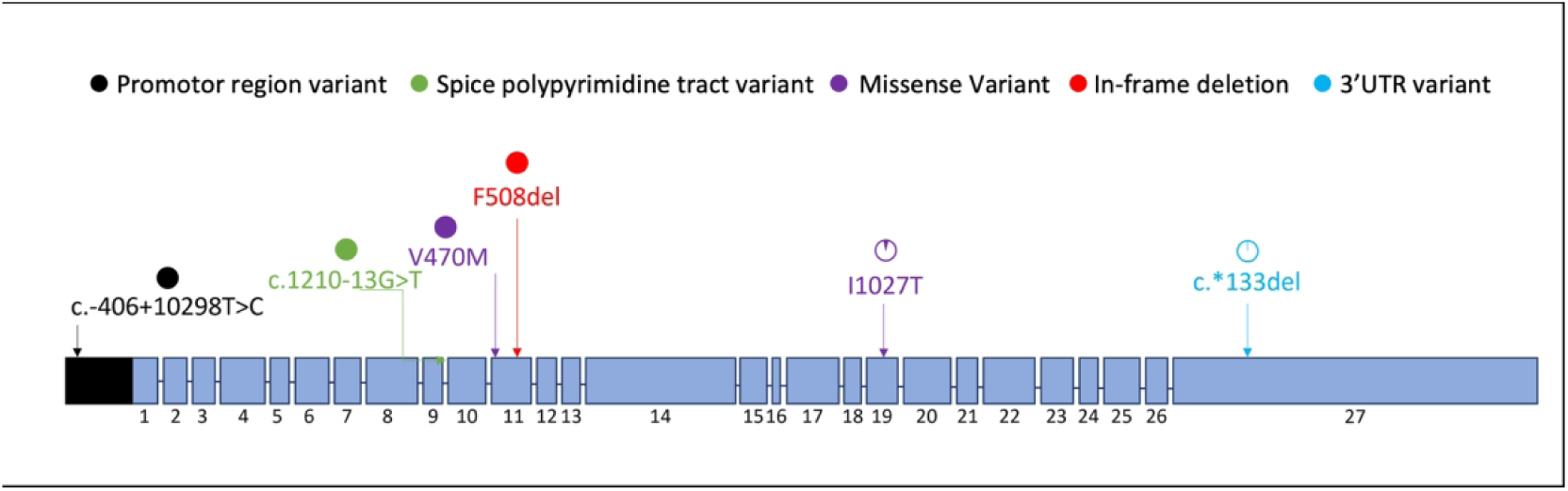
Distribution and frequency of identified variants in the CFTR gene of study participants: A schematic of non-intronic variants’ location in the CFTR gene. Each blue box represents a numbered exon, indicating its position within the gene. The frequency of each identified allele in the cohort is visualized with a progress circle, where each slice denotes a percentage of the total occurrences of that allele. Variants which were found to be homozygous in all individuals are represented by fully shaded circles. Different colours indicate the type of variant. 180×55mm (300 × 300 DPI)

### Lack of correlation between pharmacogenomics and clinical response as measured by FEV1pp and SC

Since variability in drug metabolism and transporter activity can influence drug exposure ^17,19,41^ we performed a pharmacogenetic analysis of 58 genes. To overcome the limitation of a small available sample size and distinct enzyme activity labels, we utilised descriptive heatmaps to present the relationships between specific pharmacogenetic activity and treatment efficacy (**Figure 4**). We identified considerable variability in pharmacogenes. No normal function phenotypes were observed for CYP3A5, whereas other genes displayed more variability, with CYP2C19 exhibiting phenotypes from every category.

**Figure 4:**
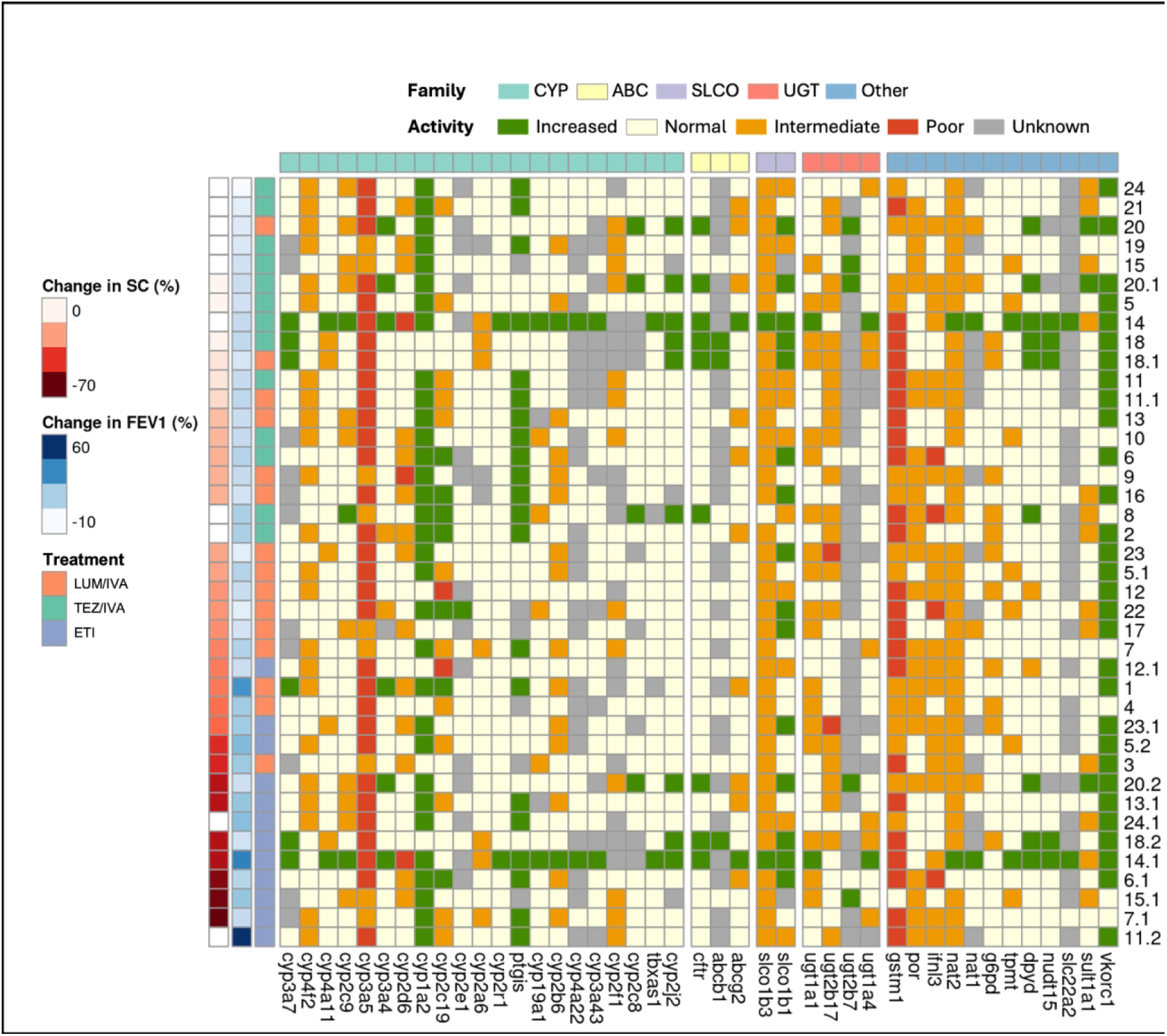
Pharmacogene profile across participants. The predicted activity levels of pharmacogenes were categorised for 58 genes from the Cytochrome P450 enzymes (CYP), ATP-binding cassette transporters (ABC), solute carrier families (SLC and SLCO) and UDP-glucuronosyltransferases (UGT). Activity levels are indicated based on their Stargazer score; increased (> 2), normal (= 2), intermediate (1 -2), poor (0 - 1) and unknown (< 0). Each column contains the predicted activity of a pharmacogene. Participants were ranked primarily by their responsiveness to treatment as measured by the relative increase in SC. FEV1pp was used to further rank participants with missing SC data. Each participant is represented once for each CFTR modulator they received. Pharmacogenes are not shown when the output for all participants was “normal” or “unknown”. SC: sweat chloride. FEV1pp: forced expiratory volume in one second, percent predicted. 181×160mm (300 × 300 DPI)

A significant degree of variability was observed in the relationships between gene function and treatment efficacy. The number of altered genes was generally consistent across individuals, except in patient #14 who possessed a high number of genes with increased function (26). Trends only emerged with individual CFTR modulators. UGT2B17 exhibited a trend towards reduced function in non-responders to LUM/IVA and ETI but not TEZ/IVA (**Figure S4A-C**). GSTM1 function was more likely to be poor in responders to TEZ/IVA and ETI but not LUM/IVA (**Figure S4A-C**). CYP2C19 and CYP2C9 activity was inversely correlated with increased clinical response to TEZ/IVA (**Figure S4B**). This was not seen for LUM/IVA and ETI (**Figure S4A, S4C**). A trend towards increased activity of PTGIS in participants with the greatest response to ETI was demonstrated (**Figure S4C**).

### Heterogenous *in vitro* functional response to CFTR modulators as measured by ΔIsc in differentiated-HNEC cultures

Differentiated-HNEC cultures created for each participant displayed pseudostratified epithelium with functional maturity, evident by mean ciliary beat frequency of 7.49 Hz and transepithelial electrical resistance (TEER) of 432 Ω.cm^2^ (**Figure S5)**^42^. Baseline CFTR activity (ΔIsc) measured in participants differentiated-HNEC cultures was less than 10% of wild type (WT) reference values (Mean 1.56%, 95% CI -0.21-3.33%) (**Figure S6**).

When participants differentiated-HNEC cultures were treated with the CFTR modulators corresponding to those received as part of their routine CF care (**Table S2**), the mean increase in ΔI_sc_ was 17.31% of WT (95% CI 9.36 – 25.26) for LUM/IVA, 11.69% of WT (95% CI 3.66 – 19.73) for TEZ/IVA and 64.28% of WT (95%CI 33.26 – 95.30) for ETI. Individual ΔIsc results for each participant highlight the heterogeneity in response to CFTR modulators between participants, independent of which modulator was used (**Figure 5B and 5C**).

**Figure 5:**
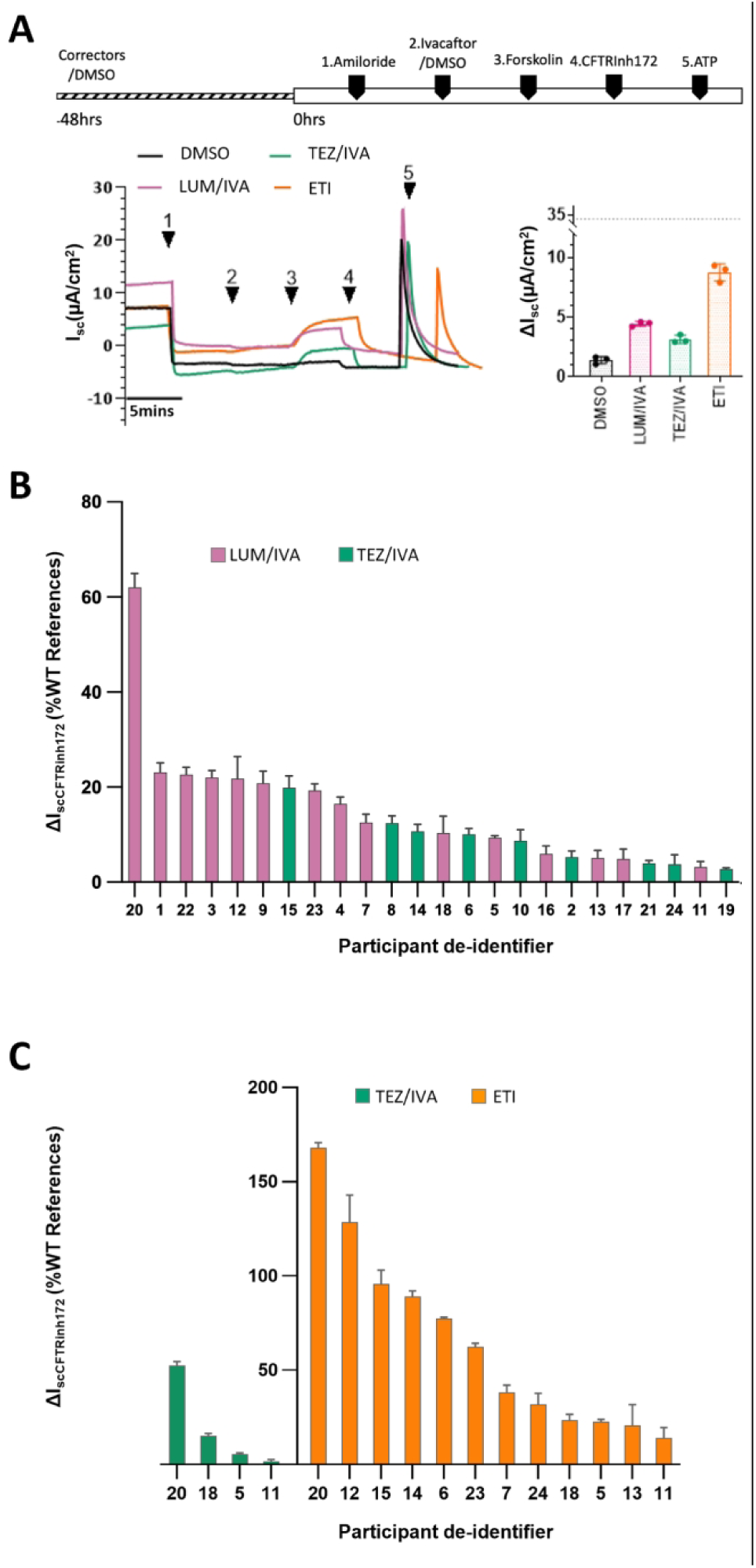
Heterogenous *in vitro* functional response to CFTR modulators. **A) Representative recordings and results from an individual participant.** Changes in short-circuit current (ΔIsc) in differentiated human nasal epithelial cell (HNEC) culture from study participant number five. Cells were either untreated (0.01% DMSO vehicle) or pre-treated with a corrector(s) (Lumacaftor (LUM), tezacaftor (TEZ:T) or elexacaftor (E) plus TEZ for 48 hours). Functional CFTR expression was then measured by sequentially adding 100 μM apical amiloride (1. Amiloride), 0.01% apical DMSO vehicle control or 10 μM apical VX-770 (ivacaftor (IVA;I )), followed by 10 μM basal forskolin (3. Forskolin), 30 μM apical CFTR inhibitor (4. CFTRinh172), and 100 μM apical ATP (5. ATP). The basolateral-to-apical chloride gradient was used to measure functional CFTR activity. The i*n vitro* ΔIsc results for participant number five are displayed on the bar chart, with the dotted line indicating the wild-type reference level for our lab (as reported in Wong et al. 2022^42^). CFTR activity is expressed as the inhibition of Isc by CFTRinh172 after activation by forskolin. Each dot represents an independent HNEC culture. Data are presented as mean ± SEM. **B) ΔIsc in differentiated HNEC cultures to first modulator treatment.** Bar chart showing ΔIsc (% of wild-type reference) in participants differentiated HNEC cultures in response to their first CFTR modulator treatment. CFTR activity is expressed as the inhibition of Isc by CFTRinh172 after activation by forskolin. Modulator response is calculated by subtracting baseline CFTR activity (DMSO) from modulator-treated results and reported as a percentage of normal (wild type). **C) ΔIsc in differentiated HNEC cultures to subsequent modulator treatments.** Bar chart showing ΔIsc (% of wild-type reference) in response to a subsequent CFTR modulator regimen. CFTR activity is expressed as the inhibition of Isc by CFTRinh172 after activation by forskolin. Changes are calculated by subtracting baseline CFTR activity (DMSO) from modulator-treated results and reported as a percentage of normal (wild type). Each bar represents the mean result of three replicate cultures per participant. Error bars represent the standard error of the mean (SEM). 113×233mm (300 × 300 DPI)

### Linear relationship between *in vitro* functional response and *in vivo* clinical response to the first CFTR modulator

Multiple linear regression was conducted to examine the relationship between the *in vitro* functional response (ΔIsc) in differentiated HNEC cultures and the participant’s *in vivo* clinical response (FEV1pp, SC) to CFTR modulator. A significant linear relationship was found between participants’ ΔIsc and their change in FEV1pp, with strong evidence of an interaction with baseline FEV1pp group (R^2^ = 0.651, p = 0.001, p <0.001, **Figure 6A**). Additionally, a significant linear relationship was identified between ΔIsc and the SC response to CFTR modulator treatment (R^2^ = 0.535, p = 0.004, **Figure 6B**).

**Figure 6:**
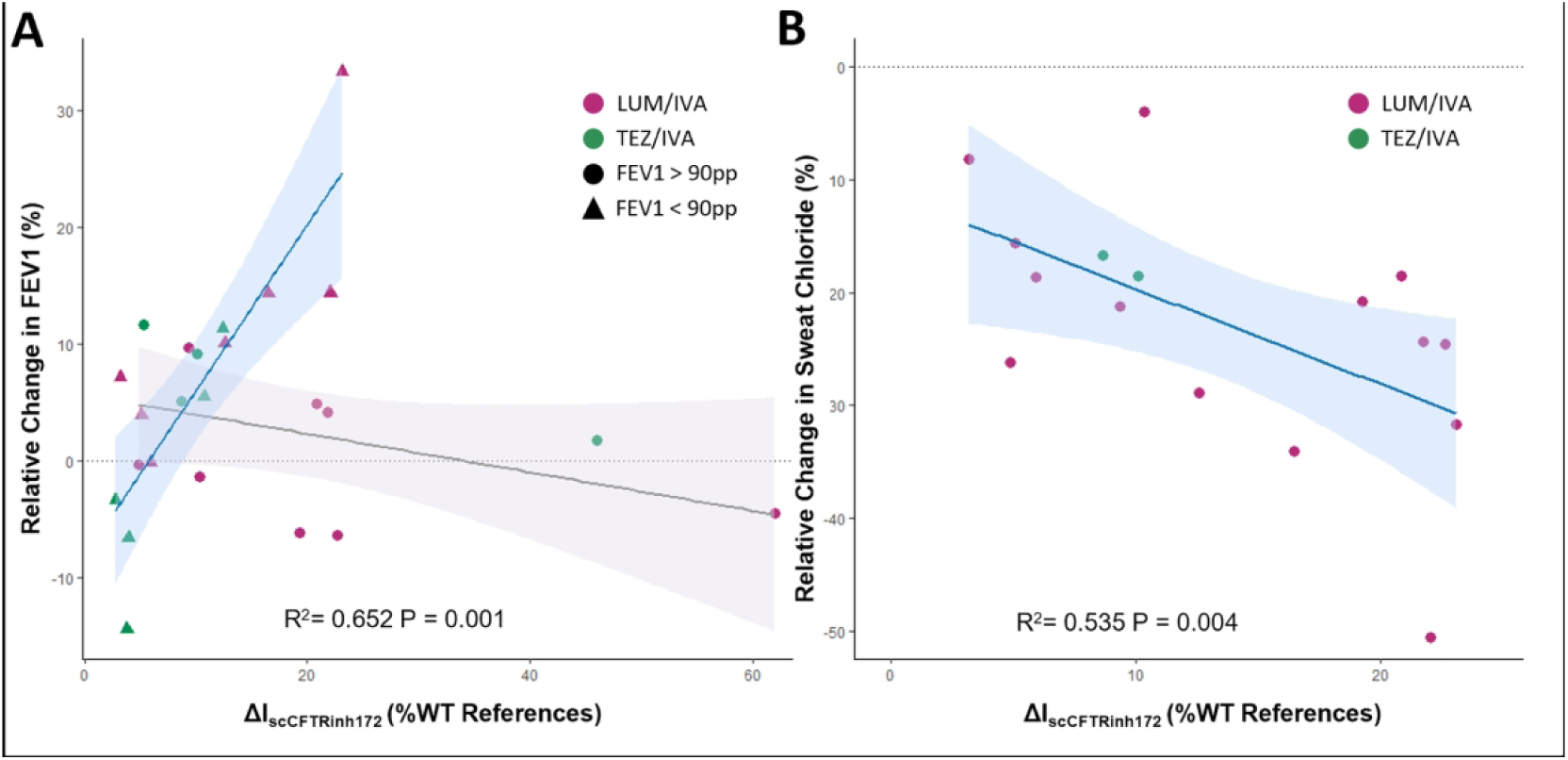
Comparison of in vitro and in vivo CFTR modulator responses. **A) FEV1pp scatterplot.** Relative change in FEV1pp is plotted against the change in short circuit current (ΔIsc) in differentiated-HNEC cultures in response to CFTR Inh172, as percentage of the wild type reference response. Each point represents an individual participant, denoted by dots (n = 12) indicating baseline FEV1pp above 90 or triangles (n = 12) indicating baseline FEV1 less than 90. Data are analysed using multiple linear regression. Separate regression lines are shown for participants with baseline FEV1 above and below 90pp. The colour of each point represents the specific CFTR modulator treatment administered. **B) SC scatterplot**. Relative change in SC is plotted against ΔIsc in differentiated-HNEC cultures in response to CFTR Inh172, as a percentage of the wild type reference response. Each dot represents a participant (n = 16). Data are analysed using multiple linear regression. The colour of each point represents the specific CFTR modulator treatment administered. The shaded area indicates the 95% confidence interval (CI) of the regression line. FEV1pp: Forced Expiratory Volume in 1 second, percent predicted. SC: Sweat Chloride. 177×85mm (300 × 300 DPI)

### Predictive capacity of *in vitro* functional response for the *in vivo* clinical response to the first CFTR modulator

To assess the capacity of differentiated-HNEC cultures to identify a pre-defined clinically significant response (FEV1pp increase > 5 percentage points, SC decrease > 20mmol/L) to treatment, we used receiver operated characteristic (ROC) curves (**Figure 7**). When examining the data as a whole, curves incorporating a decrease in SC demonstrated high predictive accuracy (AUC 0.88 (95% CI; 0.71, 1.00, and AUC 0.77 (95% CI; 0.53, 1.00), **Figure 7A**). Notably, when stratifying the data based on baseline FEV1pp being above or below 90, significant predicative accuracy was only observed in the group with a baseline FEV1pp below 90 (AUC 1.00 (95% CI; 1.00, 1.00) for both FEV1pp response and combined FEV1pp and SC response, although we note the group size was small (**Figure 7B and 7C**).

**Figure 7:**
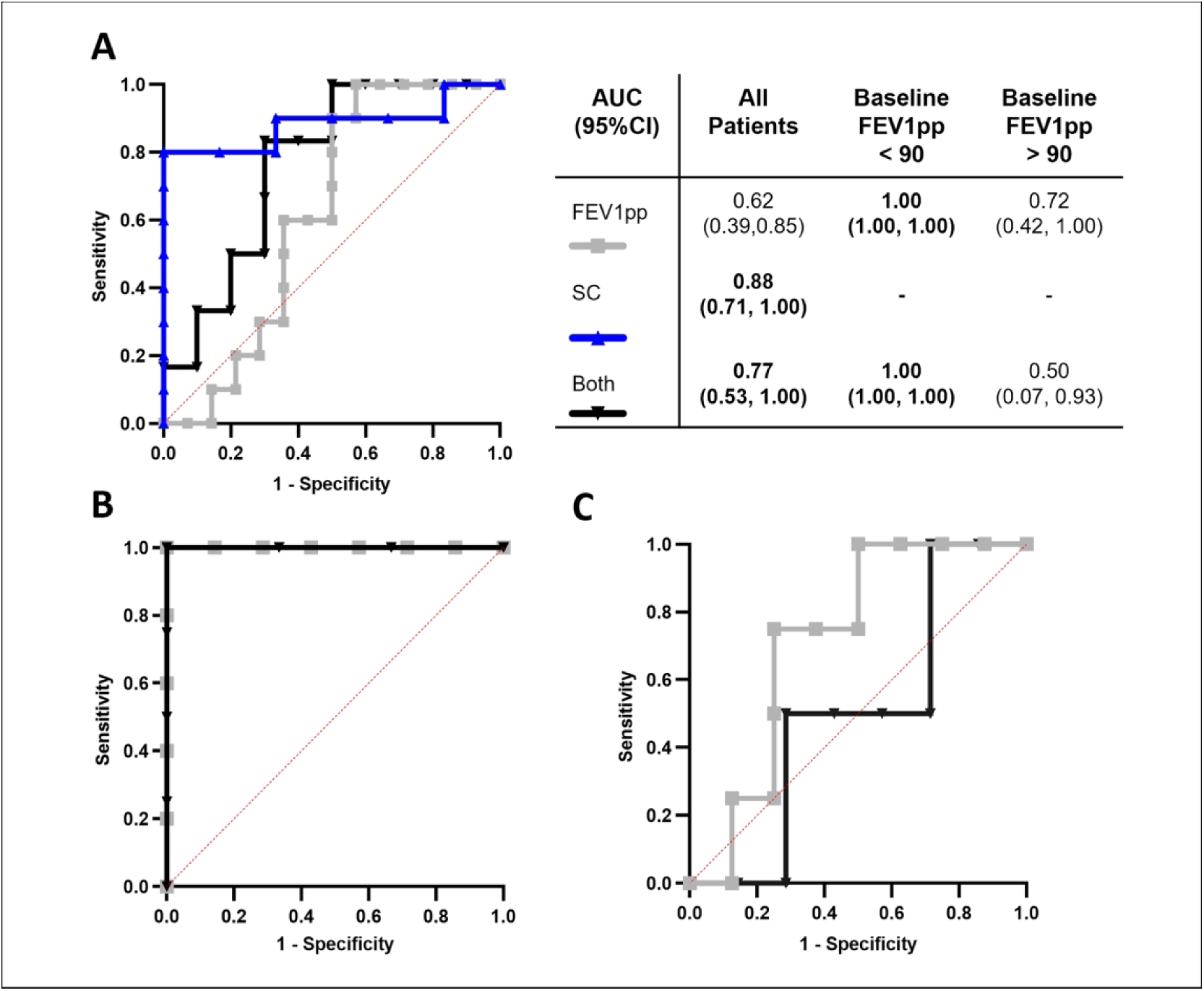
Receiver operator characteristic (ROC) curves of in vitro drug effect predicting *in vivo* clinical response. **A) ROC curves for all participants.** The predictive value of *in vitro* measurements for overall clinical responses. The grey curve represents FEV1pp (n = 24), while the blue curve represents SC (n = 16). The black curve shows the predictive value of a significant change in both FEV1pp and SC (n = 16). The diagonal red line represents the line of no discrimination. The table on the right provides the area under the curve (AUC) values with their 95% confidence intervals for different participant groups. Statistically significant results are highlighted in bold. **B) ROC curves for participants with baseline FEV1pp under 90.** The grey curve represents FEV1pp (n = 12), and the black curve shows the predictive value of a change in both FEV1pp and SC (n = 7) **C) ROC curves for participants with baseline FEV1pp above 90.** The grey curve represents FEV1pp (n = 12), and the black curve shows the predictive value of a change in both FEV1pp and SC (n = 9). Clinical Response Criteria: Increase in FEV1pp was considered positive when FEV1pp increased by at least 5 percentage points. A decrease in SC was considered positive when SC was reduced by at least 20mmol/L. FEV1pp: Forced Expiratory Volume in 1 second, percent predicted. SC: Sweat Chloride. 178×146mm (300 × 300 DPI)

### Confirmation of trends with exploratory analysis of subsequent modulator treatments

To further investigate the consistency of the differentiated-HNEC culture responses with *in vivo* clinical outcomes, we conducted exploratory analysis on participants who underwent changes in their CFTR modulator treatments. This analysis included 40 paired *in vivo-in vitro* results (**Table 1**, **Figure 8A**). Using generalised estimating equations (*see material and methods for details*), we found strong evidence that the ΔIsc predicted the change in FEV1pp (p = 0.006, **Table S4A**). Consistent with the primary analysis, baseline FEV1pp group significantly influenced the change in FEV1pp (p = 0.028, **Figure 8B, Table S4B**). We additionally observed that the choice of CFTR modulator significantly affects the change in FEV1pp (p = 0.044, **Table S4B**), and the relationship between ΔIsc and change in FEV1pp varied by CFTR modulator (p = 0.030, **Figure S7A**). Sensitivity analysis, after removing two outliers identified by checking normality of residuals after linear fit (P.d #1 and #11), showed strong evidence that ΔIsc predicts change in FEV1pp and confirmed an interaction with baseline FEV1pp group (p < 0.001, p = 0.001, **Figure S7B, Table S4B**).

**Figure 8:**
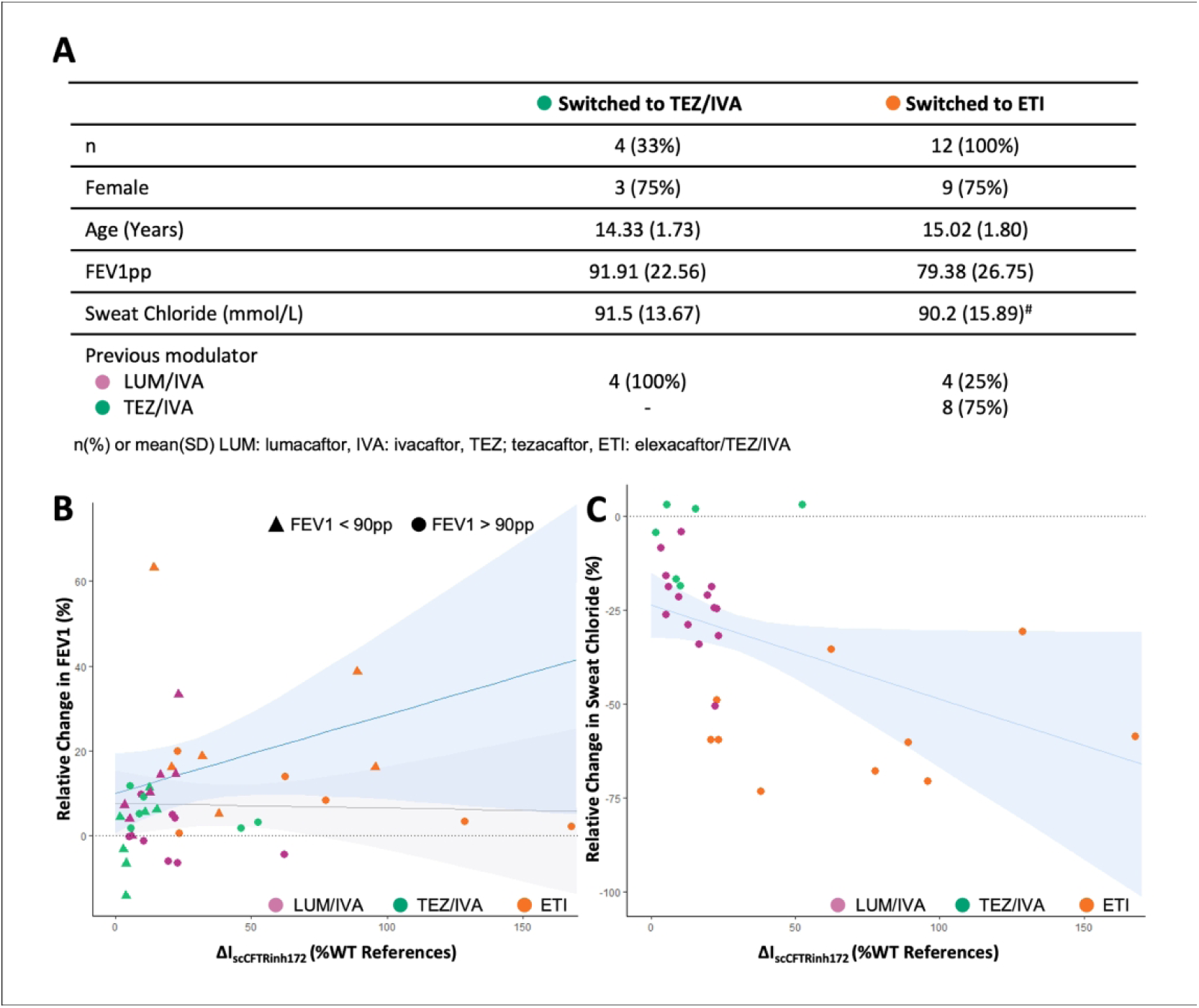
Exploratory analysis of relationship between CFTR drug response *in vitro* and *in vivo*, including subsequent modulator treatments. **A) Patient Characteristics.** The table summarises the characteristics of participants who switched to either tezacaftor/ivacaftor (TEZ/IVA) or elexacaftor/TEZ/IVA(ETI) treatments. Values for FEV1pp and SC are recorded immediately before switching treatments. Symbol (#) denotes three participants did not have measurements before switching to ETI. **B) FEV1pp vs. CFTR activity (ΔIsc) in differentiated HNEC cultures.** Plots of relative change in FEV1pp vs ΔIsc in response to CFTRinh172 treatment. Separate regression lines are shown for participants with baseline FEV1 above and below 90pp. The shaded area indicates the 95% confidence interval (CI). Each point (dot/triangle) represents a single comparison (n = 40). The colour of each point represents the specific CFTR modulator treatment administered **C) SC vs. CFTR activity (ΔIsc) in differentiated-HNEC cultures.** Plots of change in SC vs ΔIsc in response to CFTRinh172 treatment. The shaded area indicates the 95% confidence interval (CI). Each point represents a single comparison (n = 30 due to missing SC data for nine participants (**Table S2**)). The colour of each point represents the specific CFTR modulator treatment administered. FEV1pp: Forced Expiratory Volume in 1 second, percent predicted. SC: Sweat Chloride. 180×151mm (300 × 300 DPI)

Finally, we explored the relationship between ΔIsc and change in SC (with 30 out of 40 comparisons available). Although the initial analysis did not find a significant relationship between the ΔIsc and SC measurements (p = 0.441, **Figure 8C**), further analysis indicated that the trend varied significantly by CFTR modulator (p < 0.0001, **Figure S7C**). After excluding the CFTR modulator variable from the statistical model and including age and gender variables only, we found strong evidence that the ΔIsc predicted change in SC (p = 0.008, **Table S4C, Figure S7D**).

## DISCUSSION

Our findings support the importance of personalised medicine in CF, where genetic and phenotypic heterogeneity profoundly influence treatment outcomes ^1,22^. We demonstrated the predictive value of differentiated-HNEC cultures, particularly in identifying significant reduction in SC levels (> 20mmol/L) and improvement in FEV1pp (> 5 percentage points) in participants with baseline FEV1pp below 90.

The variable response to CFTR modulators, highlighted by our secondary analysis involving subsequent treatments, emphasises the complexities in drug efficacy ^7,8^. Extending our analysis to include subsequent modulator treatments for 12 participants, we found that the predictive capability of differentiated-HNEC cultures remained robust, though variation across different CFTR modulators was observed in SC response. In addition, we sought to ascertain if the heterogenous response to CFTR modulator treatments, could be explained by additional variants. Specific allele combinations in *cis*, such as L467F;F508del have been shown to negatively impact the efficacy of CFTR modulators ^20,21,43–45^. In our analysis, no individuals with the L467F variant were identified. However, we found 231 variants beyond F508del, with participants being homozygous for 142 of these variants. We identified no correlation between CFTR promoter variants during and treatment response. The remaining variants, primarily located in deep intronic regions and unique to individual participants, did not provide clear insights into their relationship with modulator response.

Three participants carried the complex allele I1027T;F508del (haplotype AB). The I1027T variant has shown response to IVA and ETI modulators in heterologous immortalised cell lines and the I1027T;F508del-CFTR variant demonstrated significant response to LUM/IVA treatment ^20,46^. Although this allele did not compromise the response to ETI in our participants, only one of the three showed clinical improvement with TEZ/IVA treatment. Reports of PwCF carrying the I1027T;F508del are sparse but suggest no impairment in CFTR modulator response ^14,20,40,47^. The detection of I1027T;F508del in three participants, without uncovering novel complex alleles associated with modulator response, suggests that the heterogeneity in drug response extends beyond CFTR genotype. In addition, CFTR interacts with a complex network of proteins, known as the CFTR interactome, which regulates its stability, expression, and function ^48^. Variations in these interacting proteins, such as chaperones, degradation pathways, and ion channel regulators, can affect how CFTR responds to treatment. This complexity necessitates a multifaceted approach to treatment personalisation, incorporating both genetic and functional data to guide modulator selection and dosage adjustment ^17,20^.

We identified participants with variable response to different modulators (**Figure S2**). We did not identify any specific haplotype that was associated with variable response. Our data suggest that CFTR genotype alone does not fully predict drug response due to the influence of various factors, including genetic modifiers, and individual variations in drug metabolism ^49–51^. We extended our genomic analysis to predict the activity of participants drug metabolising enzymes (pharmacogenes). Although our analysis of these pharmacogenes was limited by the small sample size available, some trends were observed. Differences in the predicted activity of CYP450 enzymes involved in CFTR modulator metabolism did not correlate with changes in FEV1pp and SC. Furthermore, trends differed between CFTR modulators. As such, the inconsistency in relationships between pharmacogene status and treatment efficacy complicates the identification of reliable genetic biomarkers for predicting positive drug responses across CFTR therapies. Future studies should strive to investigate pharmacogenes in larger participant cohorts.

Our study contributes valuable insights into the utility of differentiated-HNEC cultures in paediatric CF treatment, particularly for those homozygous for the F508del*-*CFTR variant. These models demonstrate their applicability across various CFTR modulators ^31,34,36^, highlighting their potential in aiding the selection of appropriate modulator therapy in a clinical setting, tailored to the individuals’ genetic and phenotypic profile. Differentiated-HNEC cultures could be used to identify non responders to prevent exposure to side effects and unnecessary cost to patients and/or health systems by the use of an ineffective treatment ^52^. However, the observed plateau effect in clinical responses (FEV1pp or SC), despite maximal correction of CFTR function as achieved by ETI, necessitates additional consideration of the CF treatment paradigm. This effect, suggestive of a limit to the clinical benefits achievable with modulator therapy, was noted in our analysis of ETI data, resonating with recent findings that proposes a ceiling treatment efficacy at approximately 30% CFTR protein function correction^34^. This phenomenon indicates the need for alternative clinical measures or endpoints in clinical trials and care strategies, particularly for potent modulators like ETI, where traditional non-sensitive endpoints, such as spirometry, may not capture the full spectrum of patient benefits.

Addressing the limitations of our study, including the pragmatic challenges of data collection in a paediatric clinical setting, we acknowledge the need for comprehensive and systematic approaches to data gathering. This is particularly critical in the context of SC measurements, given the intraindividual variability of SC levels (up to 18 mmol/L) and the sensitivity to short term adherence to CFTR modulator ^53,54^. Future research should strive for completeness and accuracy in clinical data to better understand the nuances of modulator efficacy and patient response. Similarly, the *in vitro* studies may benefit from further standardisation, for example various concentrations of CFTR modulators are tested when used alone or in combination ^4,21,38^, which may impact the magnitude of CFTR functional response. A global consensus on the drug concentration used in experiments will be advantageous to compare data across laboratories.

## Conclusion

As more CFTR modulators become available for patients to choose from ^55^, identifying the right drug for each patient will become increasingly challenging for CF clinicians. Our study demonstrates the clinical significance of assessing response to CFTR modulators in children and adolescents with CF. Studies of PwCF with FEV1pp above 90 or those unable to perform spirometry will require alternative clinical endpoints to evaluate their lung disease response to modulator treatment. Screening our cohort did not identify any CFTR-genotypic reason for non-response to treatment. However, functional testing of patient derived differentiated-HNEC cultures may have prevented non responders from receiving ineffective treatments. Thus, a combination of genetic and functional testing may provide a more optimal approach for managing complex diseases such as CF.

## Data Availability

All data produced in the present study are available upon reasonable request to the authors or are contained in the manuscript.

## Author Contributions

Conceptualisation: SAW, AJ; Recruitment and Consent: LKF, SAW; Collection of clinical data: LKF; Culture and Electrophysiology: LKF; LC-MS-MS: EKS-F, Sequence analysis: ZC, HP; Formal analysis: LKF; Visualization: LKF, ZC; Writing—original draft: LKF, SAW; Writing—review and editing: SAW, LKF, KMA, ZC, EKS-F, HP, AJ; Resources: SAW, AJ; Supervision; SAW, AJ; Funding acquisition: SAW, AJ. All authors have read and agreed to the published version of the manuscript.

## Acknowledgments

We thank the study participants and their families for their contribution. We appreciate the assistance from Sydney Children’s Hospital, Randwick respiratory department in the organisation and collection of participant biospecimens – special thanks to Ms Roxanne Strachan, Ms Leanne Plush, Ms Amanda Thomsen and Ms Rhonda Bell. We thank Ms Nihan Turgutoglu for her role in the creation of the miCF biobank and Dr Samitha Fernando for his assistance with DNA extraction. Advice regarding the statistical analysis was provided by Dr Gordana Popovic from Stats Central, UNSW. This work was supported in part by Sydney Children’s Hospitals Foundation and an Australian National Health and Medical Research Council grant (NHMRC_APP1188987), Rebecca L. Cooper Foundation project grant, Cystic Fibrosis Australia, The David Millar Giles Innovation Grant, and Luminesce Alliance Research grants. LF is supported by the Rotary Club of Sydney Cove-Sydney Children’s Hospitals Foundation and UNSW postgraduate award scholarships. KA is supported by an Australian Government Research Training Program Scholarship. HRP is supported by the Australian National Health and Medical Research Council grant (NHMRC_APP2021172). EKS-F is supported by the National Health and Medical Research Council (Grant ID: APP1157287), Cure4CF, Cystic Fibrosis Foundation (SCHNEI24I0) and The University of Melbourne. SAW is supported by UNSW Scientia program and the Australian National Health and Medical Research Council (NHMRC_APP1188987). The initial results from this study were presented at TSANZ Christchurch, New Zealand in March 2023 and ECFS Vienna, Austria in June 2023.

## Conflicts of Interest

SAW is the recipient of a Vertex Innovation Grant (2018) and a TSANZ/Vertex Research Award (2020). EKS-F received the TSANZ/Vertex Research Award (2019). All are unrelated and outside of the scope of the submitted manuscript. AJ has received consulting fees from Vertex on projects unrelated to this study. LF has received sponsorship from Vertex to attend educational meetings unrelated to this study. HRP holds equity in sequencing platform companies used in the study. All other authors declare no conflict of interest.

## Supplementary File 1: Figures and Tables.

**Table S1:** Summary of studies undertaking comparative analysis of in *vivo in vitro* responses to CFTR modulators using differentiated-HNEC cultures.

| Publication | Participant age group | Participant genotypes | Participant modulator therapy | Results |
| --- | --- | --- | --- | --- |
| Pranke I et al, 2017 [1] | > 12 years | F508del homozygous (n = 7) | LUM/IVA | **FEV1pp**<br>$R^2 = 0.95$ $p < 0.0001$ |
|  |  | F508del/ S549N (n = 1) | IVA |  |
| McGarry M et al, 2017 [2] | >16 years | Various; (n = 7) | IVA | **SC**<br>Individual results reported |
|  |  | F508del/1154insTC; G542X/3849p10kbC->T; F508del/Y563N; R334W/681delC; 1717-1G->A/G85E; 1717-1G->A/A455E; F508del/A455E |  |  |
| Pranke I et al, 2019 [3] | > 12 years | F508del homozygous (n = 16) | LUM/IVA | **FEV1pp**<br>$r = 0.735$ $p = 0.004$ |
| Debley JS et al, 2020 [4] | > 18 Years | R117H; G551D (n = 7) | IVA | **FEV1pp**<br>$r = 0.71$ $R^2 = 0.51$ $P = 0.05$ |
| | | | | **SC**<br>$r = 0.85$ $R^2 = 0.72$ $P < 0.02$ |
| Dreano E et al, 2023[5] | 11 Children (median 12 years)<br>28 adults (> 18 years) | Various; (n = 39) | ETI | **FEV1pp**<br>$\rho = 0.787$, $p < 0.0001$.<br>A non-linear relationship was observed overall. In samples with $\leq 12\%$, correction, the relationship between CFTR correction level and FEV1 % pred improvement was linear ($\rho = 0.707$, $p < 0.0001$) |
| | | I601F/621+2T>G; R347P/N1303K; G85E/G85E (n = 2); S492F/R1066C; G85E/2622+1G>A; R74W;V201M;D1270N/711+1G>T; M1101K/3396delC; S977F;TG12T5/S977F;TG12T5; H1085R/N1303K (n=2); I507del/711+1G>T; N1303K/G542X; N1303K/N1303K; N1303K/CFTRdele3-10,14b-16; N1303K/R1162X; 1717-1G>A/3659delC; 1717-1G>A/2183AA>G; R1066C/E585X; Q552P/711+1G>T; G542X/1717-1G>A; 357delC/357delC; CFTRdele19/CFTRdele19; G542X/4271delC; 2183AA>G/2183AA>G; M1T/M1T; W1282X/1078delT; 4096-3C>G/4096-3C>G; R334W/M1T; 4374+1G>A/4374+1G>A; Q493X/2183AA>G; 711+1G>T/E1104X; 1525-1G>A/1525-1G>A; 711+1G>T/711+1G>T; R553X/1717-1G>A; L558S/2183AA>G; W1063X/W1063X; 2789+5G>A/K710X; 1525-1G>A/1525-1G>A | | **SC**<br>$\rho = -0.687$, $p < 0.0001$<br>A non-linear relationship was observed overall. In samples with $< 10\%$, correction, the relationship between CFTR correction level and SC improvement was linear ($\rho = -0.486$, $p = 0.009$). |

FEV1pp: Forced Expiratory Volume in 1 second, Percent Predicted. SC: Sweat Chloride
1. Pranke IM, Hatton A, Simonin J, et al. Correction of CFTR function in nasal epithelial cells from cystic fibrosis patients predicts improvement of respiratory function by CFTR modulators. Scientific Reports; 2017
2. McGarry ME, Illek B, Ly NP, et al. In vivo and in vitro ivacaftor response in cystic fibrosis patients with residual CFTR function: N-of-1 studies. Pediatr Pulmonol; 2017
3. Pranke I, Hatton A, Masson A, et al. Might brushed nasal cells be a surrogate for CFTR modulator clinical response? Am J Respir Crit Care Med American Thoracic Society; 2019
4. Debley JS, Barrow KA, Rich LM, et al. Correlation between ivacaftor-induced CFTR activation in airway epithelial cells and improved lung function: A proof-of-concept study. Ann Am Thorac Soc American Thoracic Society; 2020
5. Dreano E, Burgel PR, Hatton A, et al. Theratyping cystic fibrosis patients to guide elexacaftor/tezacaftor/ivacaftor out-of-label prescription. Eur Respir J; 2023

**Figure S1:**
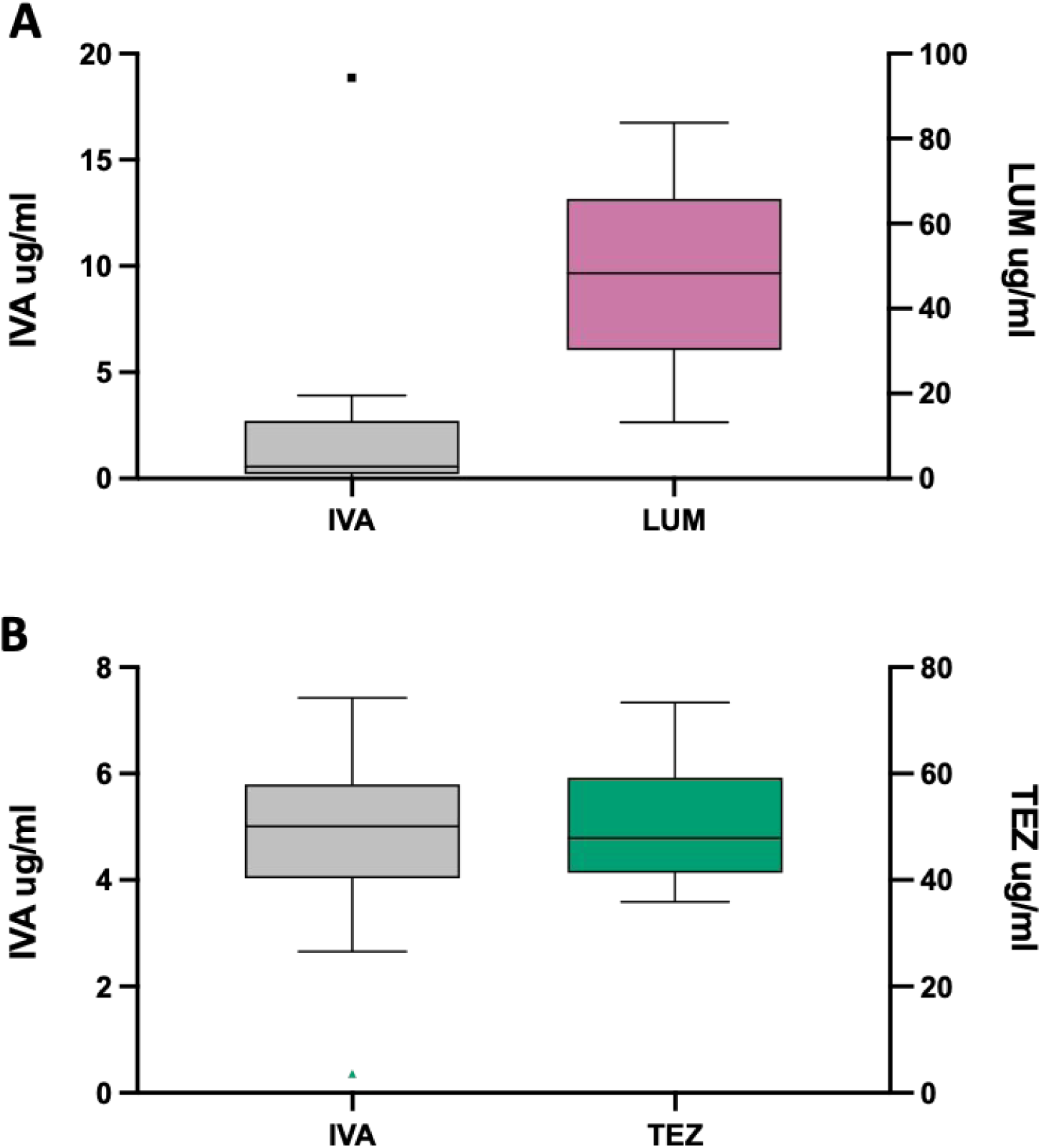
CFTR modulator levels from participants measured by LC-MS. CFTR modulator levels measured in opportunistically obtained blood samples from 14 study participants. Three participants had repeat samples from multiple time points. **A)** Ivacaftor (IVA) and lumacaftor (LUM) levels for participants on LUM/IVA (n = 7). **B)** IVA and Tezacaftor (TEZ) levels (n=7). IVA levels are plotted on the left y axis and LUM or TEZ drug levels are plotted on the right y axis. The centre line of each box blot denotes the median value. The box denotes the 25th and 75th percentiles of the dataset. The whiskers are as per Tukey formula. Any values beyond these upper and lower bounds are considered outliers and shown as a single point.

**Figure S2:**
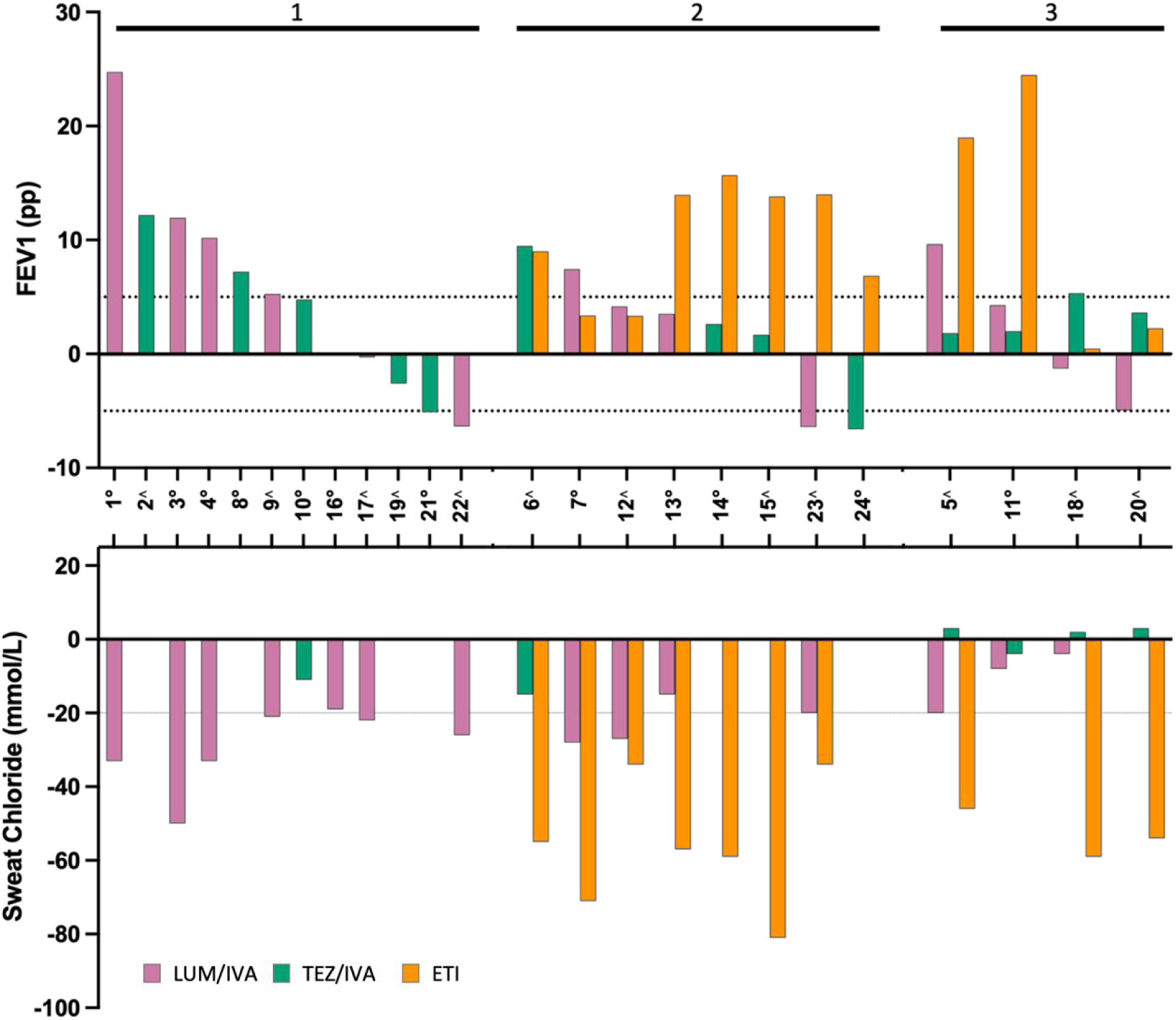
Heterogenous i*n vivo* clinical response to CFTR modulators. Waterfall plots illustrate the absolute change in participant outcomes after treatment with CFTR modulators. Results are displayed with data grouped per participant. Participants are then grouped by the number of modulator treatments received (1, 2 or 3). Absolute change in FEV1pp following treatment with lumacaftor/ivacaftor (LUM/IVA), tezacaftor/ivacaftor (TEZ/IVA) or elexacaftor/ tezacaftor/ivacaftor (ETI) is displayed on the top graph with the corresponding participant’s absolute change in SC level displayed on the lower graph. Nine participants had incomplete SC data (**Table S2**). Each bar represents an individual CFTR modulator response, with significant changes (FEV1pp change of >5 percentage points; SC decrease of > 20mmol/L) indicated by dotted lines. Participants are identified using a participant de-identifier. Symbols denote participants with a baseline FEV1pp below 90 (∘) or above 90 (∧). FEV1pp: Forced Expiratory Volume in 1 second, percent predicted. SC: Sweat Chloride.

**Figure S3:**
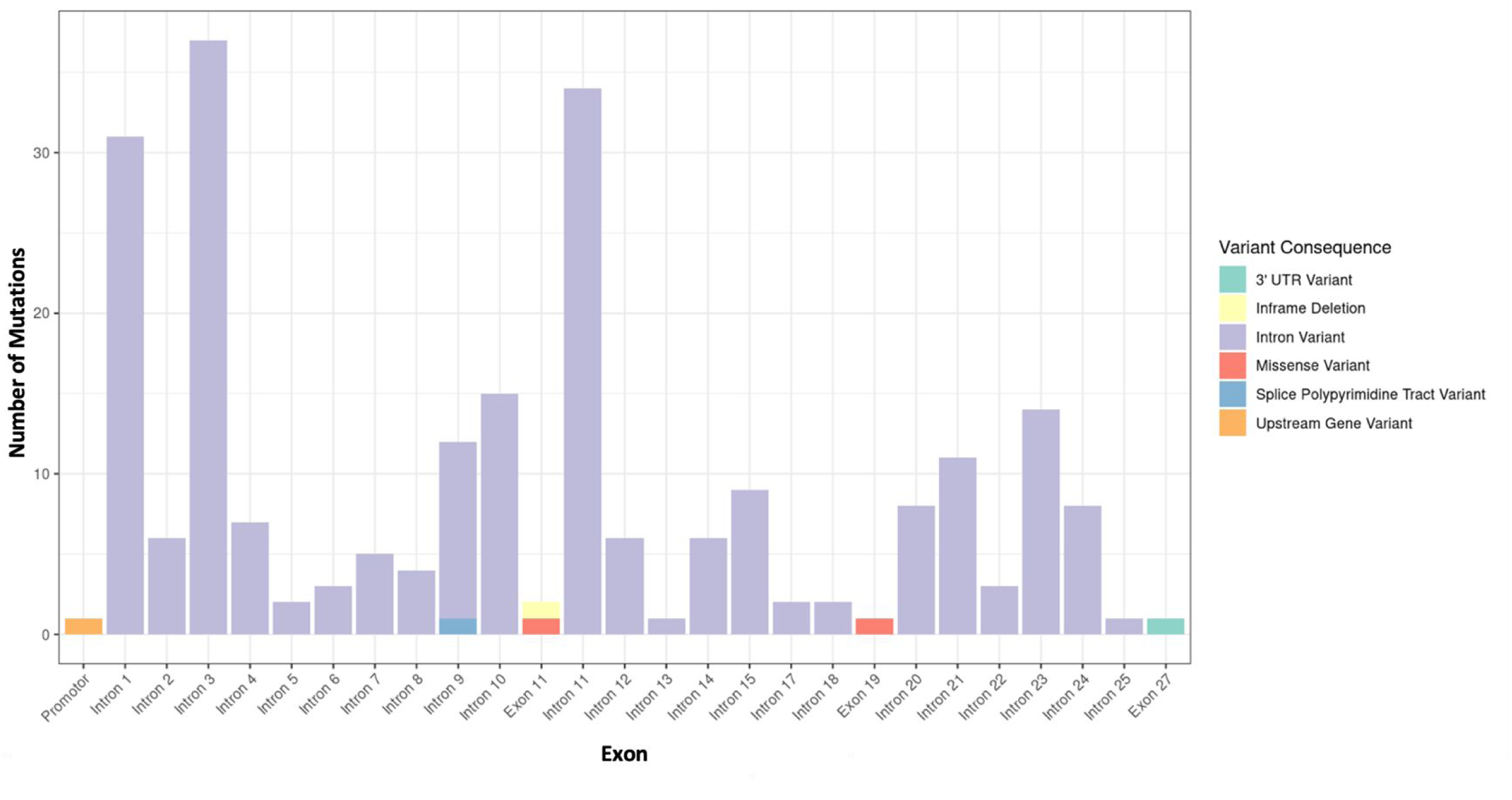
Distribution and frequency of identified variants in the CFTR gene of study participants. A histogram of the frequency and distribution of the 231 identified variants within the CFTR gene in our participants. Each bar represents one exon or intron. Only introns and exons with a frequency greater or equal to 1 are shown. Different colours highlight the type of variant.

**Figure S4:**
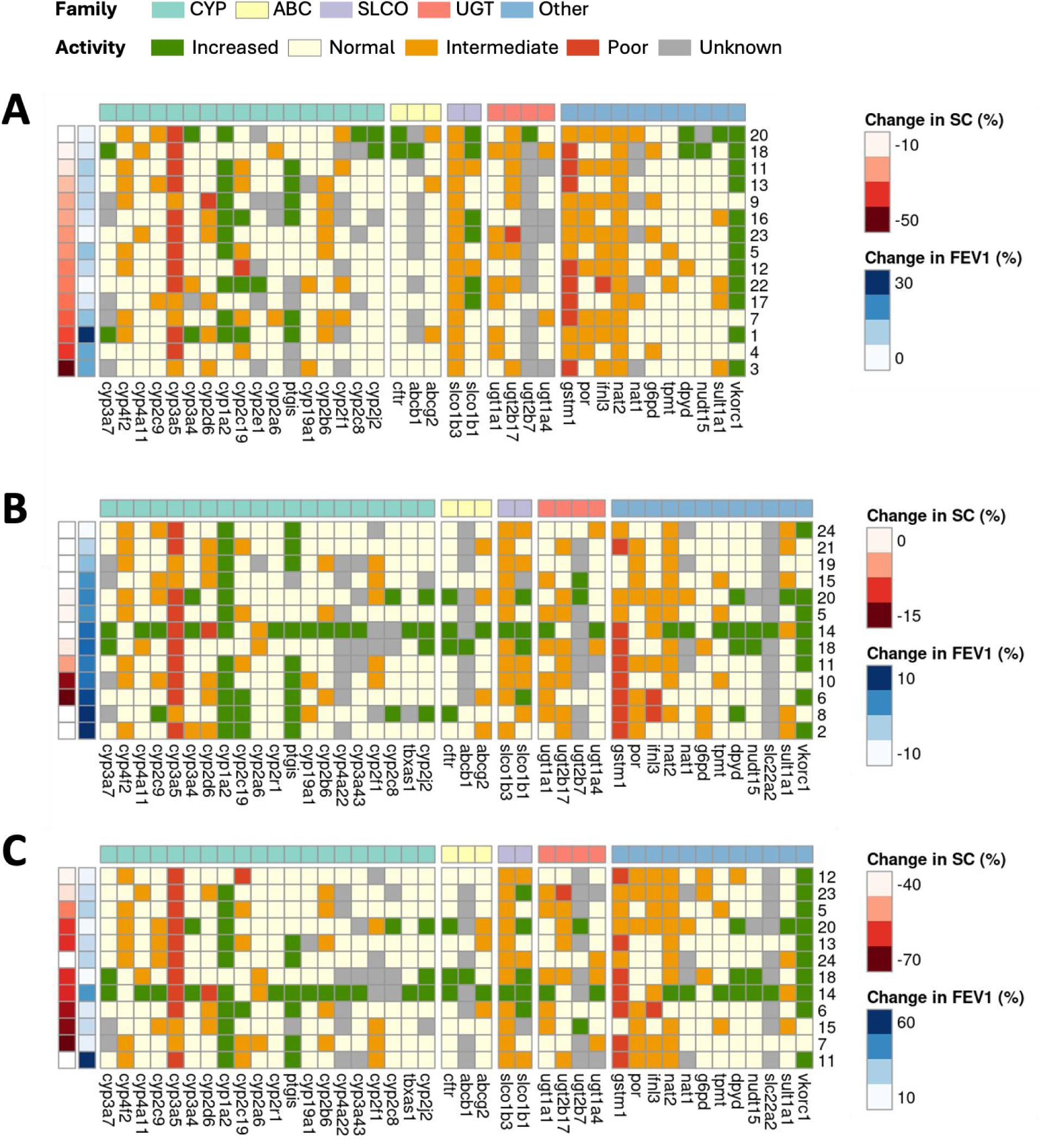
Pharmacogene profile across participants by each CFTR modulator. Pharmacogene function following treatment with **A)** Lumacaftor/Ivacaftor, **B)** Tezacaftor/Ivacaftor and **C)** Elexacaftor/Tezacaftor/Ivacaftor. The predicted activity levels of pharmacogenes were categorised for 58 genes from the Cytochrome P450 enzymes (CYP), ATP-binding cassette transporters (ABC), solute carrier families (SLC and SLCO) and UDP-glucuronosyltransferases (UGT). Activity levels are indicated based on their Stargazer score; increased (> 2), normal (= 2), intermediate (1 -2), poor (0 - 1) and unknown (< 0). Each column contains the predicted activity of a pharmacogene. Participants were ranked primarily by their responsiveness to treatment as measured by the relative increase in SC. FEV1pp was used to further rank participants with missing SC data. Each participant (n=24) is represented once per treatment and may be included up to 3 times. Pharmacogenes are not shown when the output for all participants was “normal” or “unknown”. SC: sweat chloride. FEV1pp: forced expiratory volume in one second, percent predicted.

**Supplementary table S3:**
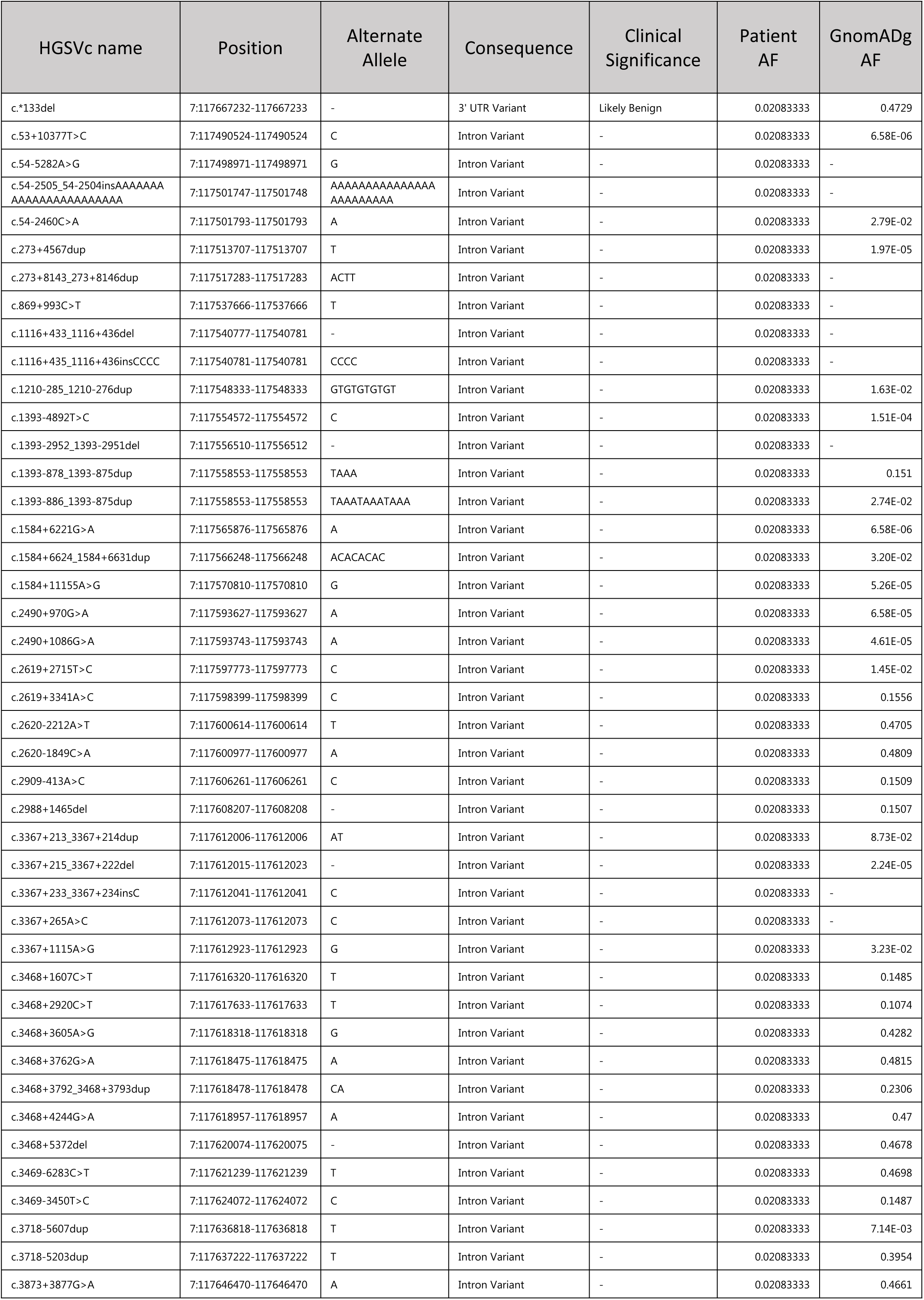
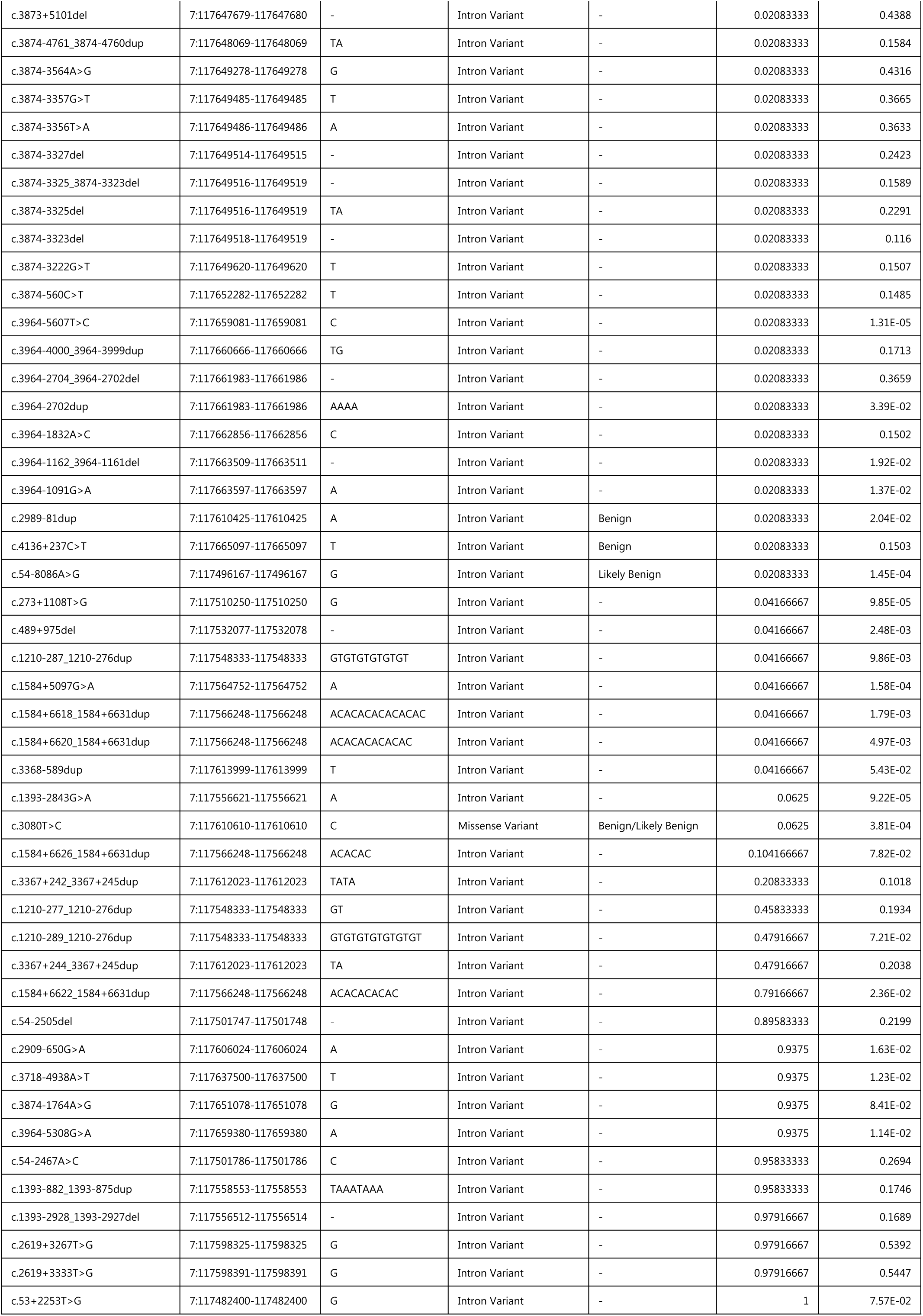
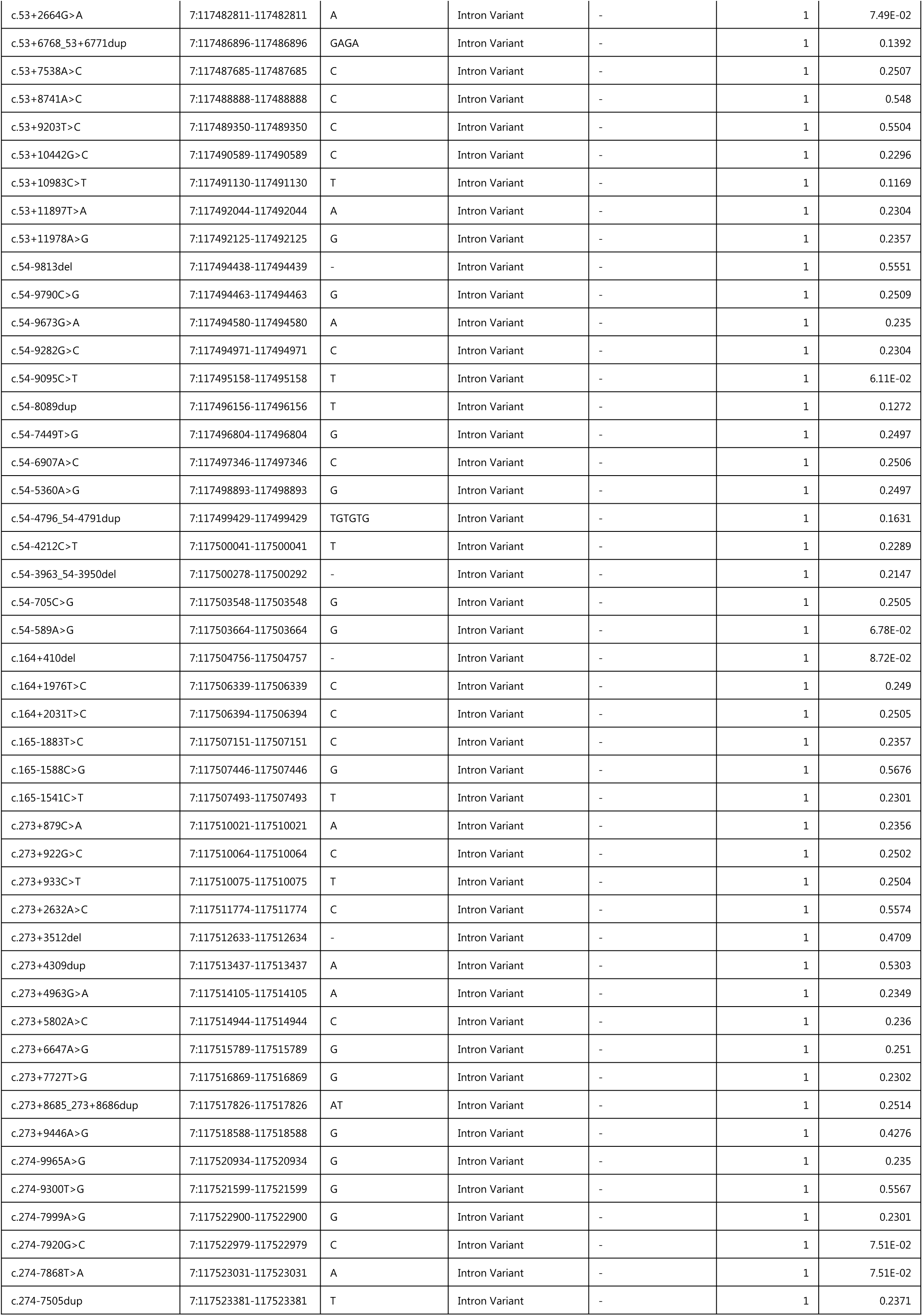
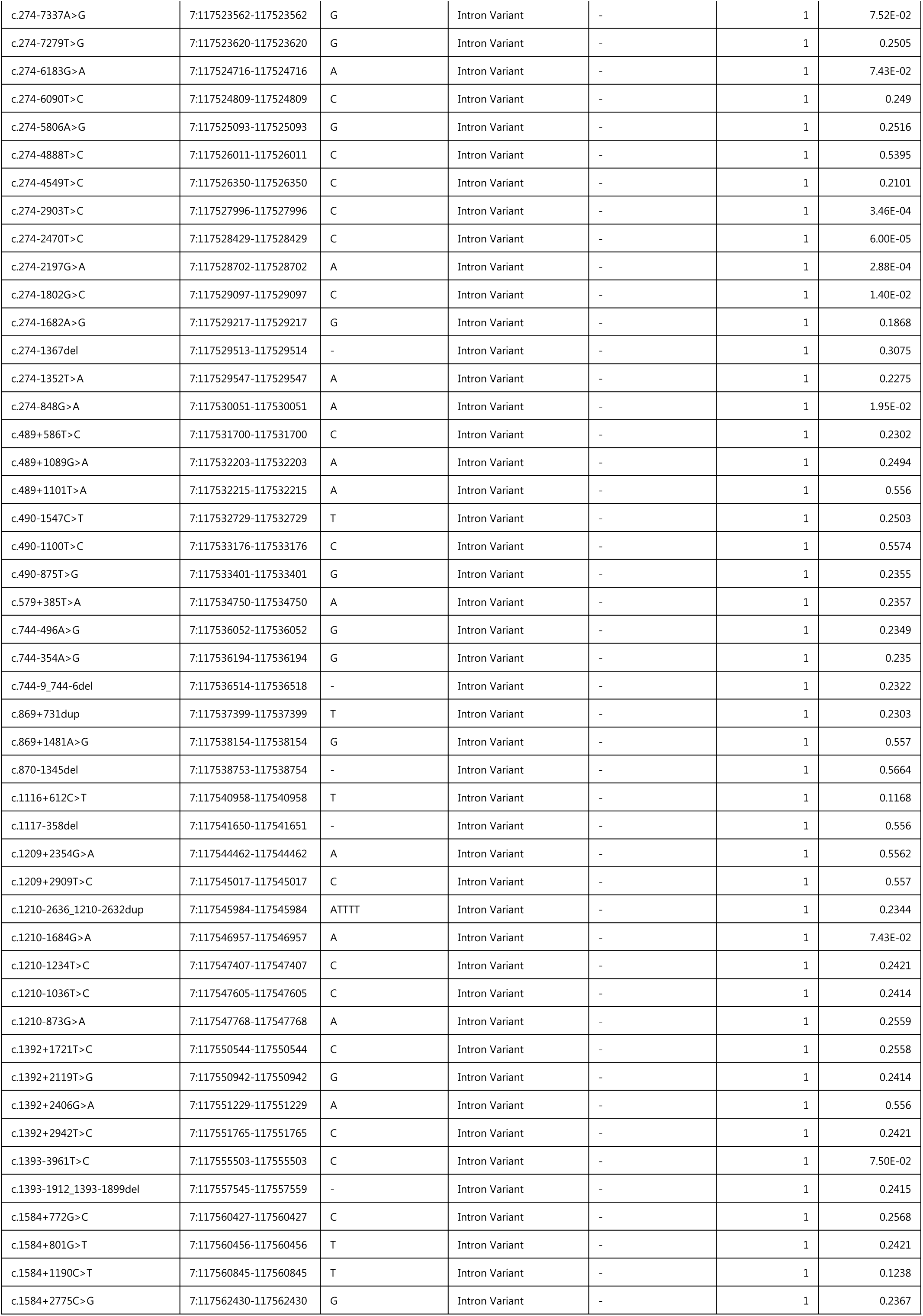
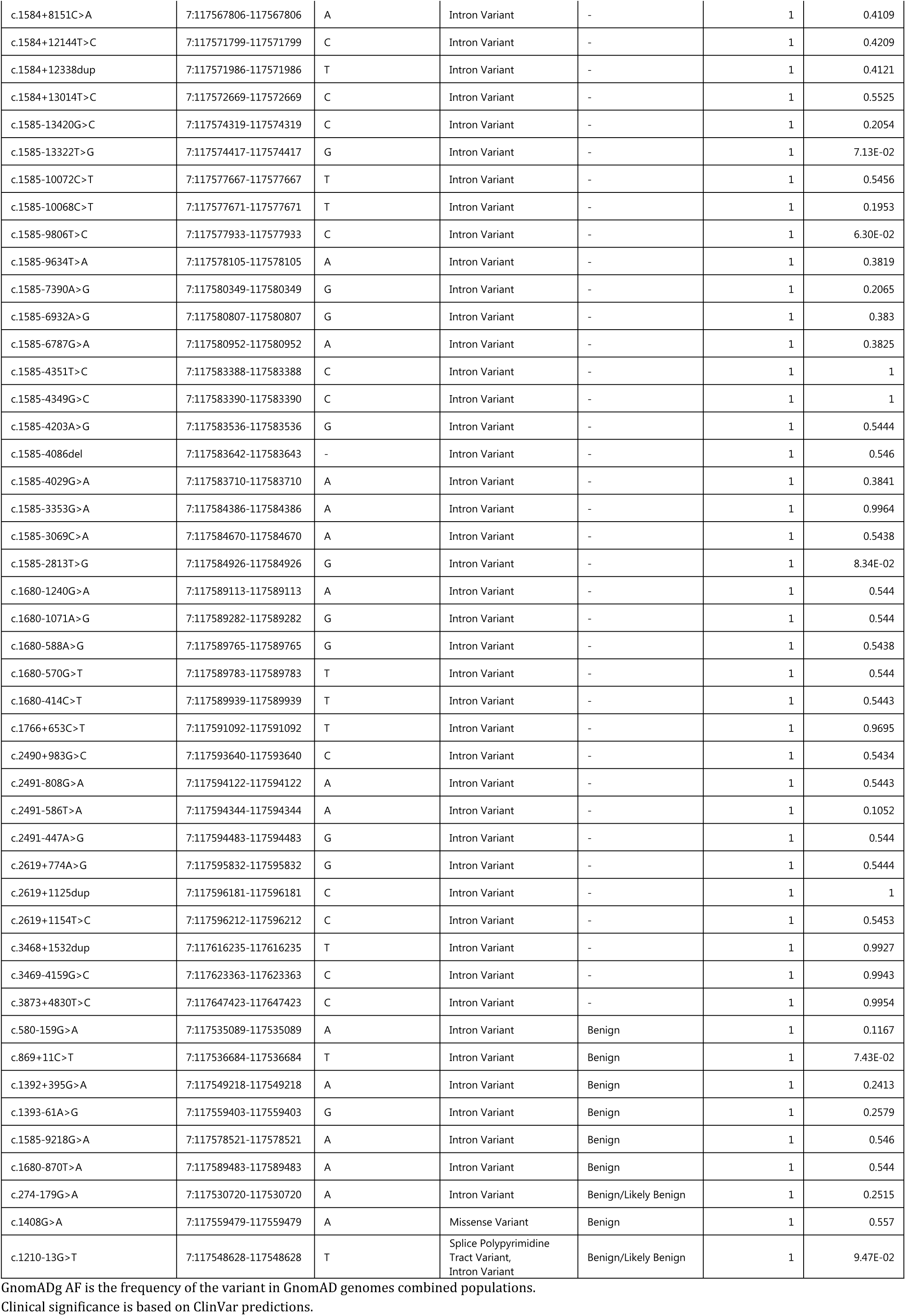
Information about the 231 additional variants detected in the cohort.

| HGSVc name | Position | Alternate Allele | Consequence | Clinical Significance | Patient AF | GnomADg AF |
| --- | --- | --- | --- | --- | --- | --- |
| c.*133del | 7:117667232-117667233 | - | 3' UTR Variant | Likely Benign | 0.02083333 | 0.4729 |
| c.53+10377T>C | 7:117490524-117490524 | C | Intron Variant | - | 0.02083333 | 6.58E-06 |
| c.54-5282A>G | 7:117498971-117498971 | G | Intron Variant | - | 0.02083333 | - |
| c.54-2505_54-2504insAAAAAA<br>AAAAAAAAAAAAAAAA | 7:117501747-117501748 | AAAAAAAAAAAAAAAA<br>AAAAAAAA | Intron Variant | - | 0.02083333 | - |
| c.54-2460C>A | 7:117501793-117501793 | A | Intron Variant | - | 0.02083333 | 2.79E-02 |
| c.273+4567dup | 7:117513707-117513707 | T | Intron Variant | - | 0.02083333 | 1.97E-05 |
| c.273+8143_273+8146dup | 7:117517283-117517283 | ACTT | Intron Variant | - | 0.02083333 | - |
| c.869+993C>T | 7:117537666-117537666 | T | Intron Variant | - | 0.02083333 | - |
| c.1116+433_1116+436del | 7:117540777-117540781 | - | Intron Variant | - | 0.02083333 | - |
| c.1116+435_1116+436insCCCC | 7:117540781-117540781 | CCCC | Intron Variant | - | 0.02083333 | - |
| c.1210-285_1210-276dup | 7:117548333-117548333 | GTGTGTGTGT | Intron Variant | - | 0.02083333 | 1.63E-02 |
| c.1393-4892T>C | 7:117554572-117554572 | C | Intron Variant | - | 0.02083333 | 1.51E-04 |
| c.1393-2952_1393-2951del | 7:117556510-117556512 | - | Intron Variant | - | 0.02083333 | - |
| c.1393-878_1393-875dup | 7:117558553-117558553 | TAAA | Intron Variant | - | 0.02083333 | 0.151 |
| c.1393-886_1393-875dup | 7:117558553-117558553 | TAAATAAATAAA | Intron Variant | - | 0.02083333 | 2.74E-02 |
| c.1584+6221G>A | 7:117565876-117565876 | A | Intron Variant | - | 0.02083333 | 6.58E-06 |
| c.1584+6624_1584+6631dup | 7:117566248-117566248 | ACACACAC | Intron Variant | - | 0.02083333 | 3.20E-02 |
| c.1584+11155A>G | 7:117570810-117570810 | G | Intron Variant | - | 0.02083333 | 5.26E-05 |
| c.2490+970G>A | 7:117593627-117593627 | A | Intron Variant | - | 0.02083333 | 6.58E-05 |
| c.2490+1086G>A | 7:117593743-117593743 | A | Intron Variant | - | 0.02083333 | 4.61E-05 |
| c.2619+2715T>C | 7:117597773-117597773 | C | Intron Variant | - | 0.02083333 | 1.45E-02 |
| c.2619+3341A>C | 7:117598399-117598399 | C | Intron Variant | - | 0.02083333 | 0.1556 |
| c.2620-2212A>T | 7:117600614-117600614 | T | Intron Variant | - | 0.02083333 | 0.4705 |
| c.2620-1849C>A | 7:117600977-117600977 | A | Intron Variant | - | 0.02083333 | 0.4809 |
| c.2909-413A>C | 7:117606261-117606261 | C | Intron Variant | - | 0.02083333 | 0.1509 |
| c.2988+1465del | 7:117608207-117608208 | - | Intron Variant | - | 0.02083333 | 0.1507 |
| c.3367+213_3367+214dup | 7:117612006-117612006 | AT | Intron Variant | - | 0.02083333 | 8.73E-02 |
| c.3367+215_3367+222del | 7:117612015-117612023 | - | Intron Variant | - | 0.02083333 | 2.24E-05 |
| c.3367+233_3367+234insC | 7:117612041-117612041 | C | Intron Variant | - | 0.02083333 | - |
| c.3367+265A>C | 7:117612073-117612073 | C | Intron Variant | - | 0.02083333 | - |
| c.3367+1115A>G | 7:117612923-117612923 | G | Intron Variant | - | 0.02083333 | 3.23E-02 |
| c.3468+1607C>T | 7:117616320-117616320 | T | Intron Variant | - | 0.02083333 | 0.1485 |
| c.3468+2920C>T | 7:117617633-117617633 | T | Intron Variant | - | 0.02083333 | 0.1074 |
| c.3468+3605A>G | 7:117618318-117618318 | G | Intron Variant | - | 0.02083333 | 0.4282 |
| c.3468+3762G>A | 7:117618475-117618475 | A | Intron Variant | - | 0.02083333 | 0.4815 |
| c.3468+3792_3468+3793dup | 7:117618478-117618478 | CA | Intron Variant | - | 0.02083333 | 0.2306 |
| c.3468+4244G>A | 7:117618957-117618957 | A | Intron Variant | - | 0.02083333 | 0.47 |
| c.3468+5372del | 7:117620074-117620075 | - | Intron Variant | - | 0.02083333 | 0.4678 |
| c.3469-6283C>T | 7:117621239-117621239 | T | Intron Variant | - | 0.02083333 | 0.4698 |
| c.3469-3450T>C | 7:117624072-117624072 | C | Intron Variant | - | 0.02083333 | 0.1487 |
| c.3718-5607dup | 7:117636818-117636818 | T | Intron Variant | - | 0.02083333 | 7.14E-03 |
| c.3718-5203dup | 7:117637222-117637222 | T | Intron Variant | - | 0.02083333 | 0.3954 |
| c.3873+3877G>A | 7:117646470-117646470 | A | Intron Variant | - | 0.02083333 | 0.4661 |

|  |  |  |  |  |  |  |  |
| --- | --- | --- | --- | --- | --- | --- | --- |
| 1 | c.3873+5101del | 7:117647679-117647680 | - | Intron Variant | - | 0.02083333 | 0.4388 |
| 2 | c.3874-4761_3874-4760dup | 7:117648069-117648069 | TA | Intron Variant | - | 0.02083333 | 0.1584 |
| 3 | c.3874-3564A>G | 7:117649278-117649278 | G | Intron Variant | - | 0.02083333 | 0.4316 |
| 4 | c.3874-3357G>T | 7:117649485-117649485 | T | Intron Variant | - | 0.02083333 | 0.3665 |
| 5 | c.3874-3356T>A | 7:117649486-117649486 | A | Intron Variant | - | 0.02083333 | 0.3633 |
| 6 | c.3874-3327del | 7:117649514-117649515 | - | Intron Variant | - | 0.02083333 | 0.2423 |
| 7 | c.3874-3325_3874-3323del | 7:117649516-117649519 | - | Intron Variant | - | 0.02083333 | 0.1589 |
| 8 | c.3874-3325del | 7:117649516-117649519 | TA | Intron Variant | - | 0.02083333 | 0.2291 |
| 9 | c.3874-3323del | 7:117649518-117649519 | - | Intron Variant | - | 0.02083333 | 0.116 |
| 10 | c.3874-3222G>T | 7:117649620-117649620 | T | Intron Variant | - | 0.02083333 | 0.1507 |
| 11 | c.3874-560C>T | 7:117652282-117652282 | T | Intron Variant | - | 0.02083333 | 0.1485 |
| 12 | c.3964-5607T>C | 7:117659081-117659081 | C | Intron Variant | - | 0.02083333 | 1.31E-05 |
| 13 | c.3964-4000_3964-3999dup | 7:117660666-117660666 | TG | Intron Variant | - | 0.02083333 | 0.1713 |
| 14 | c.3964-2704_3964-2702del | 7:117661983-117661986 | - | Intron Variant | - | 0.02083333 | 0.3659 |
| 15 | c.3964-2702dup | 7:117661983-117661986 | AAAA | Intron Variant | - | 0.02083333 | 3.39E-02 |
| 16 | c.3964-1832A>C | 7:117662856-117662856 | C | Intron Variant | - | 0.02083333 | 0.1502 |
| 17 | c.3964-1162_3964-1161del | 7:117663509-117663511 | - | Intron Variant | - | 0.02083333 | 1.92E-02 |
| 18 | c.3964-1091G>A | 7:117663597-117663597 | A | Intron Variant | - | 0.02083333 | 1.37E-02 |
| 19 | c.2989-81dup | 7:117610425-117610425 | A | Intron Variant | Benign | 0.02083333 | 2.04E-02 |
| 20 | c.4136+237C>T | 7:117665097-117665097 | T | Intron Variant | Benign | 0.02083333 | 0.1503 |
| 21 | c.54-8086A>G | 7:117496167-117496167 | G | Intron Variant | Likely Benign | 0.02083333 | 1.45E-04 |
| 22 | c.273+1108T>G | 7:117510250-117510250 | G | Intron Variant | - | 0.04166667 | 9.85E-05 |
| 23 | c.489+975del | 7:117532077-117532078 | - | Intron Variant | - | 0.04166667 | 2.48E-03 |
| 24 | c.1210-287_1210-276dup | 7:117548333-117548333 | GTGTGTGTGTGT | Intron Variant | - | 0.04166667 | 9.86E-03 |
| 25 | c.1584+5097G>A | 7:117564752-117564752 | A | Intron Variant | - | 0.04166667 | 1.58E-04 |
| 26 | c.1584+6618_1584+6631dup | 7:117566248-117566248 | ACACACACACACAC | Intron Variant | - | 0.04166667 | 1.79E-03 |
| 27 | c.1584+6620_1584+6631dup | 7:117566248-117566248 | ACACACACACAC | Intron Variant | - | 0.04166667 | 4.97E-03 |
| 28 | c.3368-589dup | 7:117613999-117613999 | T | Intron Variant | - | 0.04166667 | 5.43E-02 |
| 29 | c.1393-2843G>A | 7:117556621-117556621 | A | Intron Variant | - | 0.0625 | 9.22E-05 |
| 30 | c.3080T>C | 7:117610610-117610610 | C | Missense Variant | Benign/Likely Benign | 0.0625 | 3.81E-04 |
| 31 | c.1584+6626_1584+6631dup | 7:117566248-117566248 | ACACAC | Intron Variant | - | 0.104166667 | 7.82E-02 |
| 32 | c.3367+242_3367+245dup | 7:117612023-117612023 | TATA | Intron Variant | - | 0.20833333 | 0.1018 |
| 33 | c.1210-277_1210-276dup | 7:117548333-117548333 | GT | Intron Variant | - | 0.45833333 | 0.1934 |
| 34 | c.1210-289_1210-276dup | 7:117548333-117548333 | GTGTGTGTGTGTGT | Intron Variant | - | 0.47916667 | 7.21E-02 |
| 35 | c.3367+244_3367+245dup | 7:117612023-117612023 | TA | Intron Variant | - | 0.47916667 | 0.2038 |
| 36 | c.1584+6622_1584+6631dup | 7:117566248-117566248 | ACACACACAC | Intron Variant | - | 0.79166667 | 2.36E-02 |
| 37 | c.54-2505del | 7:117501747-117501748 | - | Intron Variant | - | 0.89583333 | 0.2199 |
| 38 | c.2909-650G>A | 7:117606024-117606024 | A | Intron Variant | - | 0.9375 | 1.63E-02 |
| 39 | c.3718-4938A>T | 7:117637500-117637500 | T | Intron Variant | - | 0.9375 | 1.23E-02 |
| 40 | c.3874-1764A>G | 7:117651078-117651078 | G | Intron Variant | - | 0.9375 | 8.41E-02 |
| 41 | c.3964-5308G>A | 7:117659380-117659380 | A | Intron Variant | - | 0.9375 | 1.14E-02 |
| 42 | c.54-2467A>C | 7:117501786-117501786 | C | Intron Variant | - | 0.95833333 | 0.2694 |
| 43 | c.1393-882_1393-875dup | 7:117558553-117558553 | TAAATAAA | Intron Variant | - | 0.95833333 | 0.1746 |
| 44 | c.1393-2928_1393-2927del | 7:117556512-117556514 | - | Intron Variant | - | 0.97916667 | 0.1689 |
| 45 | c.2619+3267T>G | 7:117598325-117598325 | G | Intron Variant | - | 0.97916667 | 0.5392 |
| 46 | c.2619+3333T>G | 7:117598391-117598391 | G | Intron Variant | - | 0.97916667 | 0.5447 |
| 47 | c.53+2253T>G | 7:117482400-117482400 | G | Intron Variant | - | 1 | 7.57E-02 |
| 1 | c.53+2664G>A | 7:117482811-117482811 | A | Intron Variant | - | 1 | 7.49E-02 |
| 2 | c.53+6768_53+6771dup | 7:117486896-117486896 | GAGA | Intron Variant | - | 1 | 0.1392 |
| 3 | c.53+7538A>C | 7:117487685-117487685 | C | Intron Variant | - | 1 | 0.2507 |
| 4 | c.53+8741A>C | 7:117488888-117488888 | C | Intron Variant | - | 1 | 0.548 |
| 5 | c.53+9203T>C | 7:117489350-117489350 | C | Intron Variant | - | 1 | 0.5504 |
| 6 | c.53+10442G>C | 7:117490589-117490589 | C | Intron Variant | - | 1 | 0.2296 |
| 7 | c.53+10983C>T | 7:117491130-117491130 | T | Intron Variant | - | 1 | 0.1169 |
| 8 | c.53+11897T>A | 7:117492044-117492044 | A | Intron Variant | - | 1 | 0.2304 |
| 9 | c.53+11978A>G | 7:117492125-117492125 | G | Intron Variant | - | 1 | 0.2357 |
| 10 | c.54-9813del | 7:117494438-117494439 | - | Intron Variant | - | 1 | 0.5551 |
| 11 | c.54-9790C>G | 7:117494463-117494463 | G | Intron Variant | - | 1 | 0.2509 |
| 12 | c.54-9673G>A | 7:117494580-117494580 | A | Intron Variant | - | 1 | 0.235 |
| 13 | c.54-9282G>C | 7:117494971-117494971 | C | Intron Variant | - | 1 | 0.2304 |
| 14 | c.54-9095C>T | 7:117495158-117495158 | T | Intron Variant | - | 1 | 6.11E-02 |
| 15 | c.54-8089dup | 7:117496156-117496156 | T | Intron Variant | - | 1 | 0.1272 |
| 16 | c.54-7449T>G | 7:117496804-117496804 | G | Intron Variant | - | 1 | 0.2497 |
| 17 | c.54-6907A>C | 7:117497346-117497346 | C | Intron Variant | - | 1 | 0.2506 |
| 18 | c.54-5360A>G | 7:117498893-117498893 | G | Intron Variant | - | 1 | 0.2497 |
| 19 | c.54-4796_54-4791dup | 7:117499429-117499429 | TGTGTG | Intron Variant | - | 1 | 0.1631 |
| 20 | c.54-4212C>T | 7:117500041-117500041 | T | Intron Variant | - | 1 | 0.2289 |
| 21 | c.54-3963_54-3950del | 7:117500278-117500292 | - | Intron Variant | - | 1 | 0.2147 |
| 22 | c.54-705C>G | 7:117503548-117503548 | G | Intron Variant | - | 1 | 0.2505 |
| 23 | c.54-589A>G | 7:117503664-117503664 | G | Intron Variant | - | 1 | 6.78E-02 |
| 24 | c.164+410del | 7:117504756-117504757 | - | Intron Variant | - | 1 | 8.72E-02 |
| 25 | c.164+1976T>C | 7:117506339-117506339 | C | Intron Variant | - | 1 | 0.249 |
| 26 | c.164+2031T>C | 7:117506394-117506394 | C | Intron Variant | - | 1 | 0.2505 |
| 27 | c.165-1883T>C | 7:117507151-117507151 | C | Intron Variant | - | 1 | 0.2357 |
| 28 | c.165-1588C>G | 7:117507446-117507446 | G | Intron Variant | - | 1 | 0.5676 |
| 29 | c.165-1541C>T | 7:117507493-117507493 | T | Intron Variant | - | 1 | 0.2301 |
| 30 | c.273+879C>A | 7:117510021-117510021 | A | Intron Variant | - | 1 | 0.2356 |
| 31 | c.273+922G>C | 7:117510064-117510064 | C | Intron Variant | - | 1 | 0.2502 |
| 32 | c.273+933C>T | 7:117510075-117510075 | T | Intron Variant | - | 1 | 0.2504 |
| 33 | c.273+2632A>C | 7:117511774-117511774 | C | Intron Variant | - | 1 | 0.5574 |
| 34 | c.273+3512del | 7:117512633-117512634 | - | Intron Variant | - | 1 | 0.4709 |
| 35 | c.273+4309dup | 7:117513437-117513437 | A | Intron Variant | - | 1 | 0.5303 |
| 36 | c.273+4963G>A | 7:117514105-117514105 | A | Intron Variant | - | 1 | 0.2349 |
| 37 | c.273+5802A>C | 7:117514944-117514944 | C | Intron Variant | - | 1 | 0.236 |
| 38 | c.273+6647A>G | 7:117515789-117515789 | G | Intron Variant | - | 1 | 0.251 |
| 39 | c.273+7727T>G | 7:117516869-117516869 | G | Intron Variant | - | 1 | 0.2302 |
| 40 | c.273+8685_273+8686dup | 7:117517826-117517826 | AT | Intron Variant | - | 1 | 0.2514 |
| 41 | c.273+9446A>G | 7:117518588-117518588 | G | Intron Variant | - | 1 | 0.4276 |
| 42 | c.274-9965A>G | 7:117520934-117520934 | G | Intron Variant | - | 1 | 0.235 |
| 43 | c.274-9300T>G | 7:117521599-117521599 | G | Intron Variant | - | 1 | 0.5567 |
| 44 | c.274-7999A>G | 7:117522900-117522900 | G | Intron Variant | - | 1 | 0.2301 |
| 45 | c.274-7920G>C | 7:117522979-117522979 | C | Intron Variant | - | 1 | 7.51E-02 |
| 46 | c.274-7868T>A | 7:117523031-117523031 | A | Intron Variant | - | 1 | 7.51E-02 |
| 47 | c.274-7505dup | 7:117523381-117523381 | T | Intron Variant | - | 1 | 0.2371 |
| 1 | c.274-7337A>G | 7:117523562-117523562 | G | Intron Variant | - | 1 | 7.52E-02 |
| 2 | c.274-7279T>G | 7:117523620-117523620 | G | Intron Variant | - | 1 | 0.2505 |
| 3 | c.274-6183G>A | 7:117524716-117524716 | A | Intron Variant | - | 1 | 7.43E-02 |
| 4 | c.274-6090T>C | 7:117524809-117524809 | C | Intron Variant | - | 1 | 0.249 |
| 5 | c.274-5806A>G | 7:117525093-117525093 | G | Intron Variant | - | 1 | 0.2516 |
| 6 | c.274-4888T>C | 7:117526011-117526011 | C | Intron Variant | - | 1 | 0.5395 |
| 7 | c.274-4549T>C | 7:117526350-117526350 | C | Intron Variant | - | 1 | 0.2101 |
| 8 | c.274-2903T>C | 7:117527996-117527996 | C | Intron Variant | - | 1 | 3.46E-04 |
| 9 | c.274-2470T>C | 7:117528429-117528429 | C | Intron Variant | - | 1 | 6.00E-05 |
| 10 | c.274-2197G>A | 7:117528702-117528702 | A | Intron Variant | - | 1 | 2.88E-04 |
| 11 | c.274-1802G>C | 7:117529097-117529097 | C | Intron Variant | - | 1 | 1.40E-02 |
| 12 | c.274-1682A>G | 7:117529217-117529217 | G | Intron Variant | - | 1 | 0.1868 |
| 13 | c.274-1367del | 7:117529513-117529514 | - | Intron Variant | - | 1 | 0.3075 |
| 14 | c.274-1352T>A | 7:117529547-117529547 | A | Intron Variant | - | 1 | 0.2275 |
| 15 | c.274-848G>A | 7:117530051-117530051 | A | Intron Variant | - | 1 | 1.95E-02 |
| 16 | c.489+586T>C | 7:117531700-117531700 | C | Intron Variant | - | 1 | 0.2302 |
| 17 | c.489+1089G>A | 7:117532203-117532203 | A | Intron Variant | - | 1 | 0.2494 |
| 18 | c.489+1101T>A | 7:117532215-117532215 | A | Intron Variant | - | 1 | 0.556 |
| 19 | c.490-1547C>T | 7:117532729-117532729 | T | Intron Variant | - | 1 | 0.2503 |
| 20 | c.490-1100T>C | 7:117533176-117533176 | C | Intron Variant | - | 1 | 0.5574 |
| 21 | c.490-875T>G | 7:117533401-117533401 | G | Intron Variant | - | 1 | 0.2355 |
| 22 | c.579+385T>A | 7:117534750-117534750 | A | Intron Variant | - | 1 | 0.2357 |
| 23 | c.744-496A>G | 7:117536052-117536052 | G | Intron Variant | - | 1 | 0.2349 |
| 24 | c.744-354A>G | 7:117536194-117536194 | G | Intron Variant | - | 1 | 0.235 |
| 25 | c.744-9_744-6del | 7:117536514-117536518 | - | Intron Variant | - | 1 | 0.2322 |
| 26 | c.869+731dup | 7:117537399-117537399 | T | Intron Variant | - | 1 | 0.2303 |
| 27 | c.869+1481A>G | 7:117538154-117538154 | G | Intron Variant | - | 1 | 0.557 |
| 28 | c.870-1345del | 7:117538753-117538754 | - | Intron Variant | - | 1 | 0.5664 |
| 29 | c.1116+612C>T | 7:117540958-117540958 | T | Intron Variant | - | 1 | 0.1168 |
| 30 | c.1117-358del | 7:117541650-117541651 | - | Intron Variant | - | 1 | 0.556 |
| 31 | c.1209+2354G>A | 7:117544462-117544462 | A | Intron Variant | - | 1 | 0.5562 |
| 32 | c.1209+2909T>C | 7:117545017-117545017 | C | Intron Variant | - | 1 | 0.557 |
| 33 | c.1210-2636_1210-2632dup | 7:117545984-117545984 | ATTTT | Intron Variant | - | 1 | 0.2344 |
| 34 | c.1210-1684G>A | 7:117546957-117546957 | A | Intron Variant | - | 1 | 7.43E-02 |
| 35 | c.1210-1234T>C | 7:117547407-117547407 | C | Intron Variant | - | 1 | 0.2421 |
| 36 | c.1210-1036T>C | 7:117547605-117547605 | C | Intron Variant | - | 1 | 0.2414 |
| 37 | c.1210-873G>A | 7:117547768-117547768 | A | Intron Variant | - | 1 | 0.2559 |
| 38 | c.1392+1721T>C | 7:117550544-117550544 | C | Intron Variant | - | 1 | 0.2558 |
| 39 | c.1392+2119T>G | 7:117550942-117550942 | G | Intron Variant | - | 1 | 0.2414 |
| 40 | c.1392+2406G>A | 7:117551229-117551229 | A | Intron Variant | - | 1 | 0.556 |
| 41 | c.1392+2942T>C | 7:117551765-117551765 | C | Intron Variant | - | 1 | 0.2421 |
| 42 | c.1393-3961T>C | 7:117555503-117555503 | C | Intron Variant | - | 1 | 7.50E-02 |
| 43 | c.1393-1912_1393-1899del | 7:117557545-117557559 | - | Intron Variant | - | 1 | 0.2415 |
| 44 | c.1584+772G>C | 7:117560427-117560427 | C | Intron Variant | - | 1 | 0.2568 |
| 45 | c.1584+801G>T | 7:117560456-117560456 | T | Intron Variant | - | 1 | 0.2421 |
| 46 | c.1584+1190C>T | 7:117560845-117560845 | T | Intron Variant | - | 1 | 0.1238 |
| 47 | c.1584+2775C>G | 7:117562430-117562430 | G | Intron Variant | - | 1 | 0.2367 |
| 1 | c.1584+8151C>A | 7:117567806-117567806 | A | Intron Variant | - | 1 | 0.4109 |
| 2 | c.1584+12144T>C | 7:117571799-117571799 | C | Intron Variant | - | 1 | 0.4209 |
| 3 | c.1584+12338dup | 7:117571986-117571986 | T | Intron Variant | - | 1 | 0.4121 |
| 4 | c.1584+13014T>C | 7:117572669-117572669 | C | Intron Variant | - | 1 | 0.5525 |
| 5 | c.1585-13420G>C | 7:117574319-117574319 | C | Intron Variant | - | 1 | 0.2054 |
| 6 | c.1585-13322T>G | 7:117574417-117574417 | G | Intron Variant | - | 1 | 7.13E-02 |
| 7 | c.1585-10072C>T | 7:117577667-117577667 | T | Intron Variant | - | 1 | 0.5456 |
| 8 | c.1585-10068C>T | 7:117577671-117577671 | T | Intron Variant | - | 1 | 0.1953 |
| 9 | c.1585-9806T>C | 7:117577933-117577933 | C | Intron Variant | - | 1 | 6.30E-02 |
| 10 | c.1585-9634T>A | 7:117578105-117578105 | A | Intron Variant | - | 1 | 0.3819 |
| 11 | c.1585-7390A>G | 7:117580349-117580349 | G | Intron Variant | - | 1 | 0.2065 |
| 12 | c.1585-6932A>G | 7:117580807-117580807 | G | Intron Variant | - | 1 | 0.383 |
| 13 | c.1585-6787G>A | 7:117580952-117580952 | A | Intron Variant | - | 1 | 0.3825 |
| 14 | c.1585-4351T>C | 7:117583388-117583388 | C | Intron Variant | - | 1 | 1 |
| 15 | c.1585-4349G>C | 7:117583390-117583390 | C | Intron Variant | - | 1 | 1 |
| 16 | c.1585-4203A>G | 7:117583536-117583536 | G | Intron Variant | - | 1 | 0.5444 |
| 17 | c.1585-4086del | 7:117583642-117583643 | - | Intron Variant | - | 1 | 0.546 |
| 18 | c.1585-4029G>A | 7:117583710-117583710 | A | Intron Variant | - | 1 | 0.3841 |
| 19 | c.1585-3353G>A | 7:117584386-117584386 | A | Intron Variant | - | 1 | 0.9964 |
| 20 | c.1585-3069C>A | 7:117584670-117584670 | A | Intron Variant | - | 1 | 0.5438 |
| 21 | c.1585-2813T>G | 7:117584926-117584926 | G | Intron Variant | - | 1 | 8.34E-02 |
| 22 | c.1680-1240G>A | 7:117589113-117589113 | A | Intron Variant | - | 1 | 0.544 |
| 23 | c.1680-1071A>G | 7:117589282-117589282 | G | Intron Variant | - | 1 | 0.544 |
| 24 | c.1680-588A>G | 7:117589765-117589765 | G | Intron Variant | - | 1 | 0.5438 |
| 25 | c.1680-570G>T | 7:117589783-117589783 | T | Intron Variant | - | 1 | 0.544 |
| 26 | c.1680-414C>T | 7:117589939-117589939 | T | Intron Variant | - | 1 | 0.5443 |
| 27 | c.1766+653C>T | 7:117591092-117591092 | T | Intron Variant | - | 1 | 0.9695 |
| 28 | c.2490+983G>C | 7:117593640-117593640 | C | Intron Variant | - | 1 | 0.5434 |
| 29 | c.2491-808G>A | 7:117594122-117594122 | A | Intron Variant | - | 1 | 0.5443 |
| 30 | c.2491-586T>A | 7:117594344-117594344 | A | Intron Variant | - | 1 | 0.1052 |
| 31 | c.2491-447A>G | 7:117594483-117594483 | G | Intron Variant | - | 1 | 0.544 |
| 32 | c.2619+774A>G | 7:117595832-117595832 | G | Intron Variant | - | 1 | 0.5444 |
| 33 | c.2619+1125dup | 7:117596181-117596181 | C | Intron Variant | - | 1 | 1 |
| 34 | c.2619+1154T>C | 7:117596212-117596212 | C | Intron Variant | - | 1 | 0.5453 |
| 35 | c.3468+1532dup | 7:117616235-117616235 | T | Intron Variant | - | 1 | 0.9927 |
| 36 | c.3469-4159G>C | 7:117623363-117623363 | C | Intron Variant | - | 1 | 0.9943 |
| 37 | c.3873+4830T>C | 7:117647423-117647423 | C | Intron Variant | - | 1 | 0.9954 |
| 38 | c.580-159G>A | 7:117535089-117535089 | A | Intron Variant | Benign | 1 | 0.1167 |
| 39 | c.869+11C>T | 7:117536684-117536684 | T | Intron Variant | Benign | 1 | 7.43E-02 |
| 40 | c.1392+395G>A | 7:117549218-117549218 | A | Intron Variant | Benign | 1 | 0.2413 |
| 41 | c.1393-61A>G | 7:117559403-117559403 | G | Intron Variant | Benign | 1 | 0.2579 |
| 42 | c.1585-9218G>A | 7:117578521-117578521 | A | Intron Variant | Benign | 1 | 0.546 |
| 43 | c.1680-870T>A | 7:117589483-117589483 | A | Intron Variant | Benign | 1 | 0.544 |
| 44 | c.274-179G>A | 7:117530720-117530720 | A | Intron Variant | Benign/Likely Benign | 1 | 0.2515 |
| 45 | c.1408G>A | 7:117559479-117559479 | A | Missense Variant | Benign | 1 | 0.557 |
| 46 | c.1210-13G>T | 7:117548628-117548628 | T | Splice Polypyrimidine Tract Variant, Intron Variant | Benign/Likely Benign | 1 | 9.47E-02 |

GnomADg AF is the frequency of the variant in GnomAD genomes combined populations. Clinical significance is based on ClinVar predictions.

**Figure S5:**
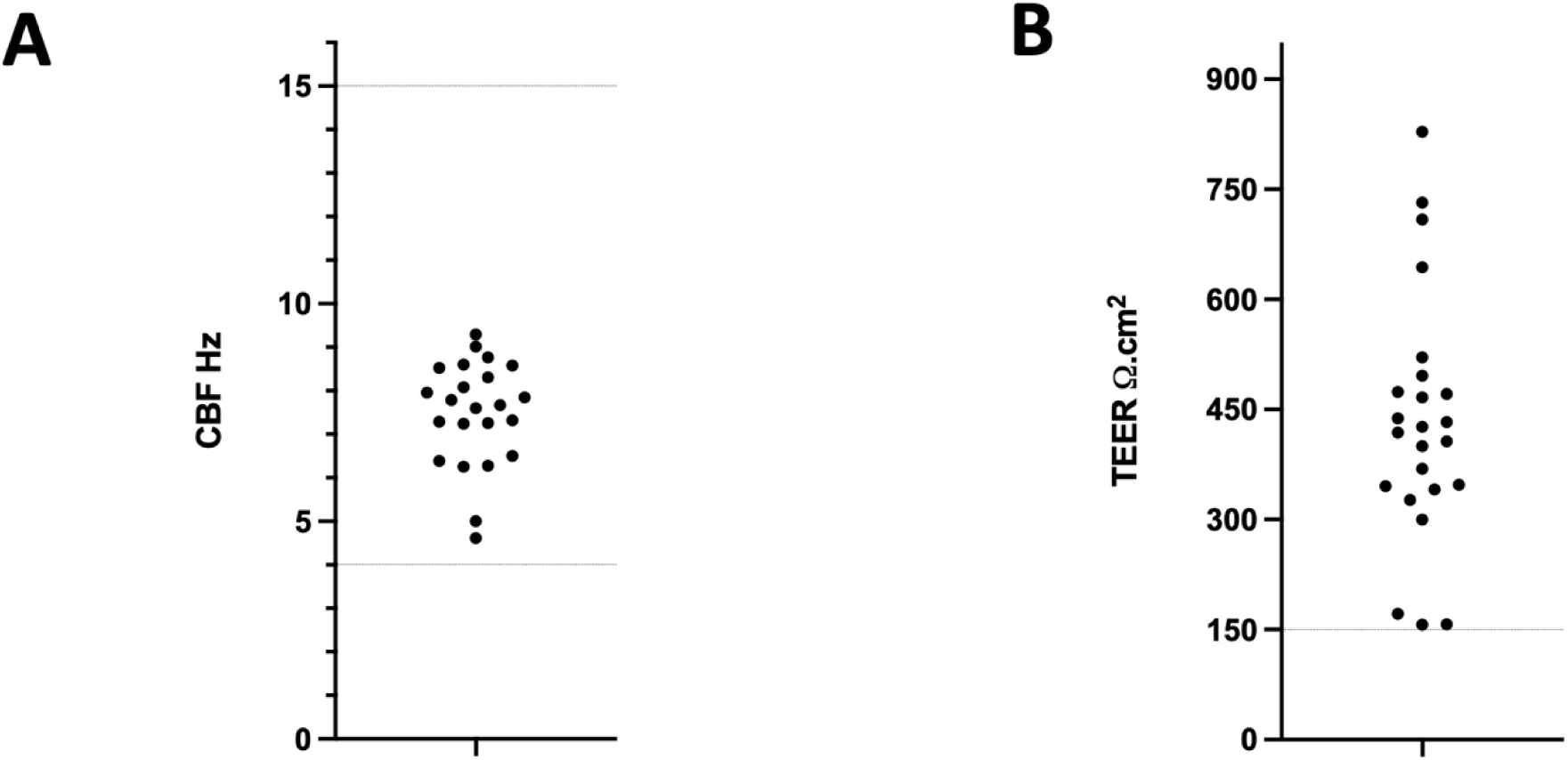
Validation of *in vitro* differentiated-HNEC cultures prior to electrophysiological assessment. A) Ciliary beat frequency (CBF). Baseline measurements of mature fully differentiated human nasal epithelial cell (HNEC) cultures prior to treatment with CFTR correctors. Each dot represents one individual (n = 23 due to failed data acquisition for one participant). Mean CBF 7.49Hz (95% CI 6.96 – 8.01). Dotted lines represent the upper and lower limits of normal values[6]. Data are represented as the mean of at least four replicate differentiated-HNEC cultures per participant. An average of six fields of view (FOV) were measured per differentiated-HNEC culture. **B) Trans epithelial electrical resistance (TEER) measurements.** TEER was measured during electrophysiological analysis of short circuit current in an Ussing chamber. Each dot represents one individual. Mean transepithelial electrical resistance (TEER) 432Ω.cm^2^ (95% CI 361 – 504). Data are represented as the mean of at least n = 6 replicate differentiated-HNEC cultures per participant. The minimal accepted value of 150Ω.cm2 is shown with a dotted line[7]. 6. Nikolaizik W, Hahn J, Bauck M, et al. Comparison of ciliary beat frequencies at different temperatures in young adults. ERJ Open Res; 2020 7. Wong SL, Awatade NT, Astore MA, *et al.* Molecular Dynamics and Theratyping in Airway and Gut Organoids Reveal R352Q-CFTR Conductance Defect. *Am J Respir Cell Mol Biol*; 2022

**Figure S6:**
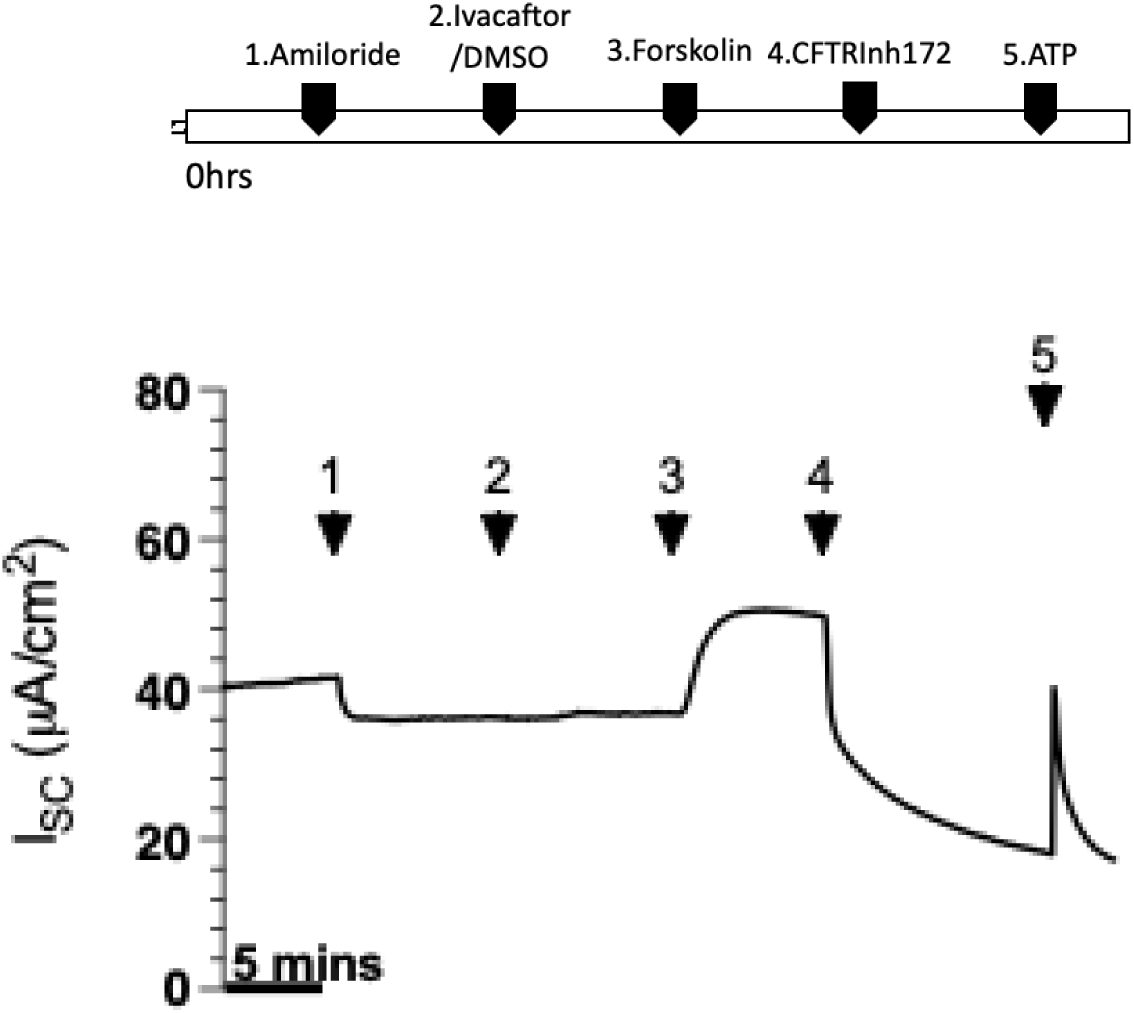
Representative wild type (WT) Ussing trace. Changes in short-circuit current (ΔIsc) in differentiated human nasal epithelial cell (HNEC) culture from WT reference (as reported in Wong et al. 2022[7]). Functional CFTR expression was measured by sequentially adding 100 μM apical amiloride (1. Amiloride), 0.01% apical DMSO vehicle control, followed by 10 μM basal forskolin (3. Forskolin), 30 μM apical CFTR inhibitor (4. CFTRinh172), and 100 μM apical ATP (5. ATP). The basolateral-to-apical chloride gradient was used to measure functional CFTR activity 7. Wong SL, Awatade NT, Astore MA, *et al.* Molecular Dynamics and Theratyping in Airway and Gut Organoids Reveal R352Q-CFTR Conductance Defect. *Am J Respir Cell Mol Biol*; 2022

**Figure S7:**
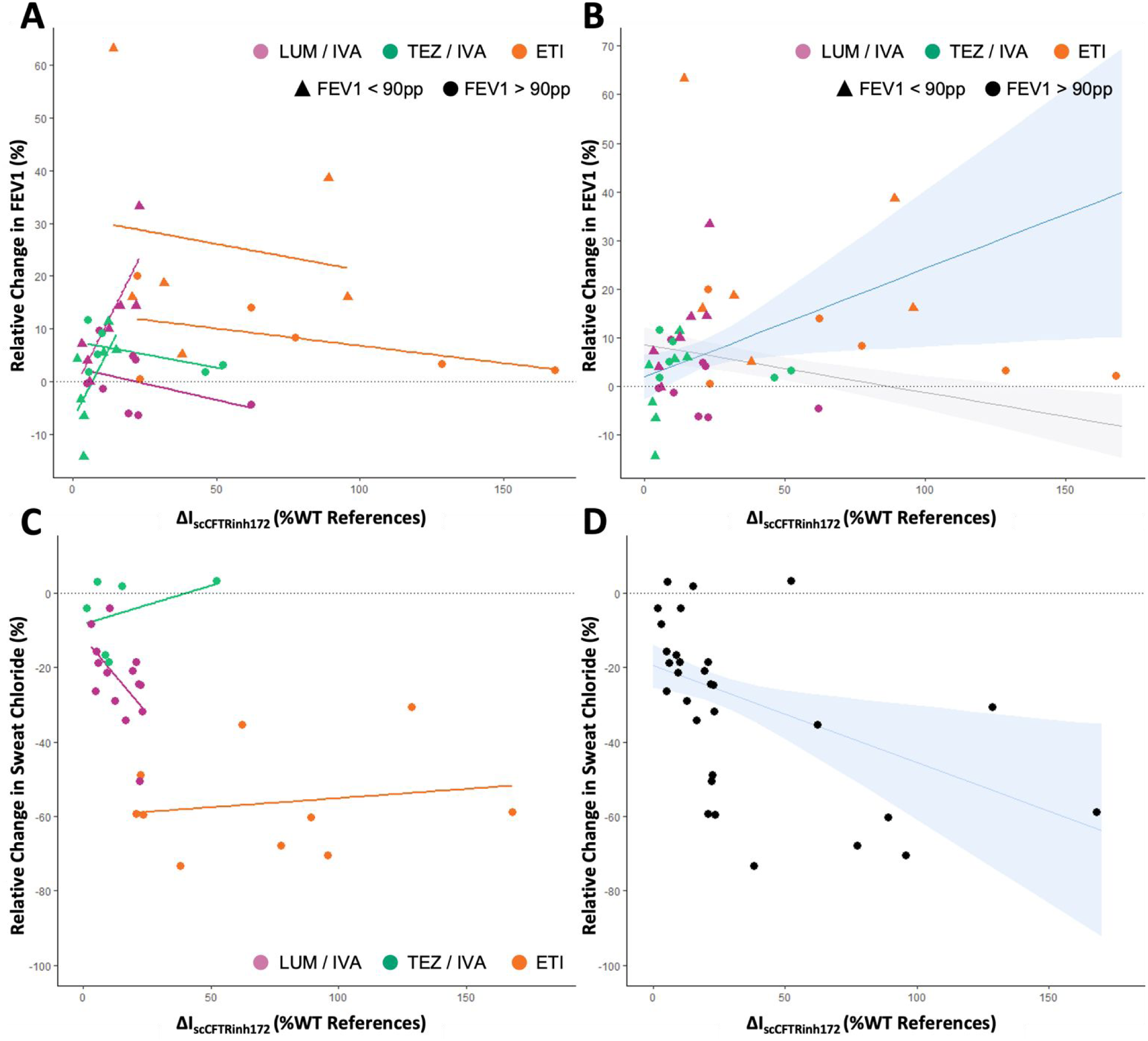
Exploring the relationship between *in vitro* functional responses to CFTR modulators by each CFTR modulator treatment and sensitivity analysis. **A) Scatterplot of FEV1pp vs. change in *in vitro* CFTR activity (ΔIsc) in differentiated-HNEC cultures.** Relative change in FEV1pp is plotted against ΔIsc in response to CFTR Inh172, as a percentage of the wild type reference response. Each point (dot/triangle) represents a single comparison (n = 40). Data are analysed using generalised estimating equations with a variance components covariance structure to account for repeated measures. Separate regression lines are shown for participants with baseline FEV1pp above and below 90 and for each CFTR modulator. The colour of each point and regression line represents the specific CFTR modulator treatment administered. **B) Scatterplot of FEV1pp vs. change in *in vitro* CFTR activity (ΔIsc) in differentiated-HNEC cultures following removal of two outliers identified during sensitivity analysis of the statistical model.** Plots of relative change in FEV1pp vs ΔIsc in response to CFTRinh172 treatment. The shaded area indicates the 95% confidence interval (CI). Each point (dot/triangle) represents a single comparison (n = 38). **C) Scatterplot of SC vs. change in *in vitro* CFTR activity (ΔIsc) in differentiated-HNEC cultures.** Relative change in SC is plotted against the ΔIsc in response to CFTR inh172, as a percentage of the wildtype reference response. Each dot represents a single comparison (n = 30). Data are analysed using generalised estimating equations with a variance components covariance structure to account for repeated measures. The colour of each point and regression line represents the specific CFTR modulator treatment administered. **D) Scatterplot of SC vs. change in *in vitro* CFTR activity (ΔIsc) in differentiated-HNEC cultures following removal of CFTR modulator treatment variable from the statistical model.** Scatterplot of relative change in SC against ΔIsc in response to CFTR Inh172, with individual comparisons represented by single dots (n=30). Data were analysed using generalised estimating equations with a variance components covariance structure to account for repeated measures. Shaded area shows 95% CI of the regression line. FEV1pp: Forced Expiratory Volume in 1 second, percent predicted. SC: Sweat Chloride.

**Table S4:**
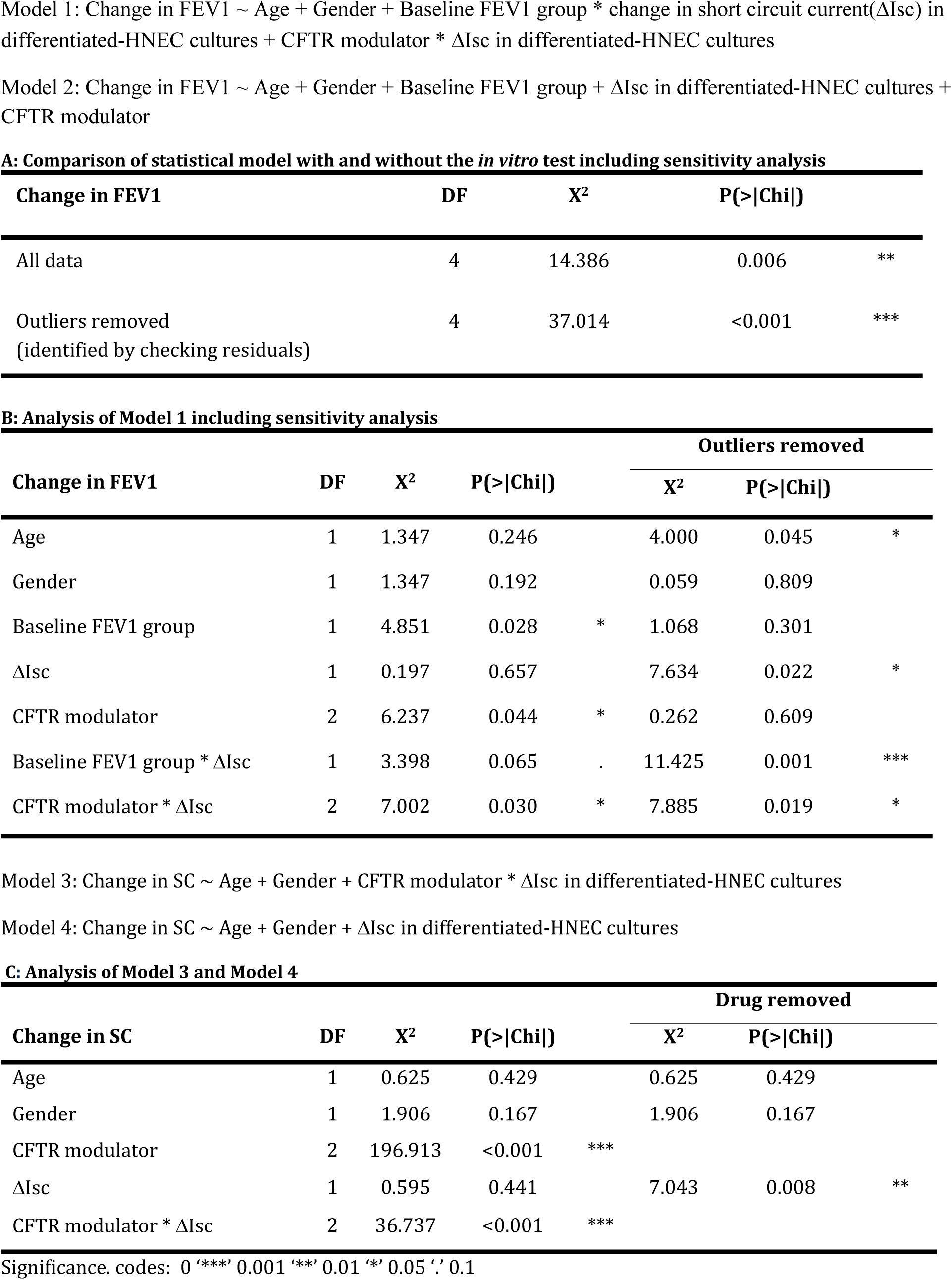
Stats results for Exploratory analysis of subsequent modulators. Analysis of 'Wald statistic' Tables

## Supplementary file #2

### Materials and Methods

#### Participants

This study was approved by the Sydney children’s Hospital Network (SCHN) Ethics Review Board (HREC/16/SCHN/120). Written informed consent was obtained from all participating subjects and/or their legal guardians. Participants were recruited from the Sydney Children’s Hospital (SCH) cystic fibrosis (CF) clinic if they; 1) had an F508del homozygous *CFTR* genotype, 2) commenced treatment with a CFTR modulator with ongoing follow up by the CF team and 3) completed spirometry testing pre and post modulator treatment according to the American Thoracic Society (ATS) criteria [1].

#### LC-MS to measure CFTR modulator drug levels

Participants with a blood sample stored in our CF biobank that was collected whilst receiving CFTR modulator treatment were identified. Samples were collected from participants opportunistically during routine collection of blood samples for clinically indicated reasons. Plasma and serum were frozen and stored at -80°C. Liquid chromatography/mass spectroscopy (LC-MS) was used to quantify ivacaftor (IVA), lumacaftor (LUM), and/or tezacaftor (TEZ) in plasma or serum as previously described [2, 3] Plasma or serum were processed with 0.1% formic acid/ACN in a 1:2 dilution ratio. Mixtures were centrifuged (10 minutes, 132,000 rpm). Supernatant were transferred into LC-MS vials. The LC-MS analysis was performed using Shimadzu LC-MS 8050 triple quadrupole mass systems.

#### *In vivo* clinical response to modulator – FEV1 percent predicted (pp)

Spirometry was performed by a respiratory scientist in an accredited respiratory laboratory as part of routine CF care [4]. Global Lung Index (GLI) references were used for all spirometry data. The clinical status of the patient was recorded at each time point to identify exacerbation events or intercurrent illnesses, as well as any dose adjustments to modulator treatments. Spirometry results obtained during exacerbations were excluded.

Baseline FEV1pp was defined as the result obtained closest to the introduction of each CFTR modulator treatment to the patient’s treatment plan. Post treatment FEV1pp was defined as the earliest FEV1pp result obtained after at least six weeks of stable CFTR modulator dosing. Six weeks was chosen to reduce the impact of the transient bronchoconstriction side effect associated with initiation of LUM/IVA treatment [5]. Due to the progressive nature of CF lung disease, when participants changed CFTR modulator treatment the FEV1pp response was calculated as the change between the FEV1pp measurement obtained immediately prior to the first dose of the subsequent CFTR modulator (subsequent baseline, **Figure M1**) and the FEV1pp result obtained after at least six weeks of stable subsequent CFTR modulator dosing.

**Figure M1:**
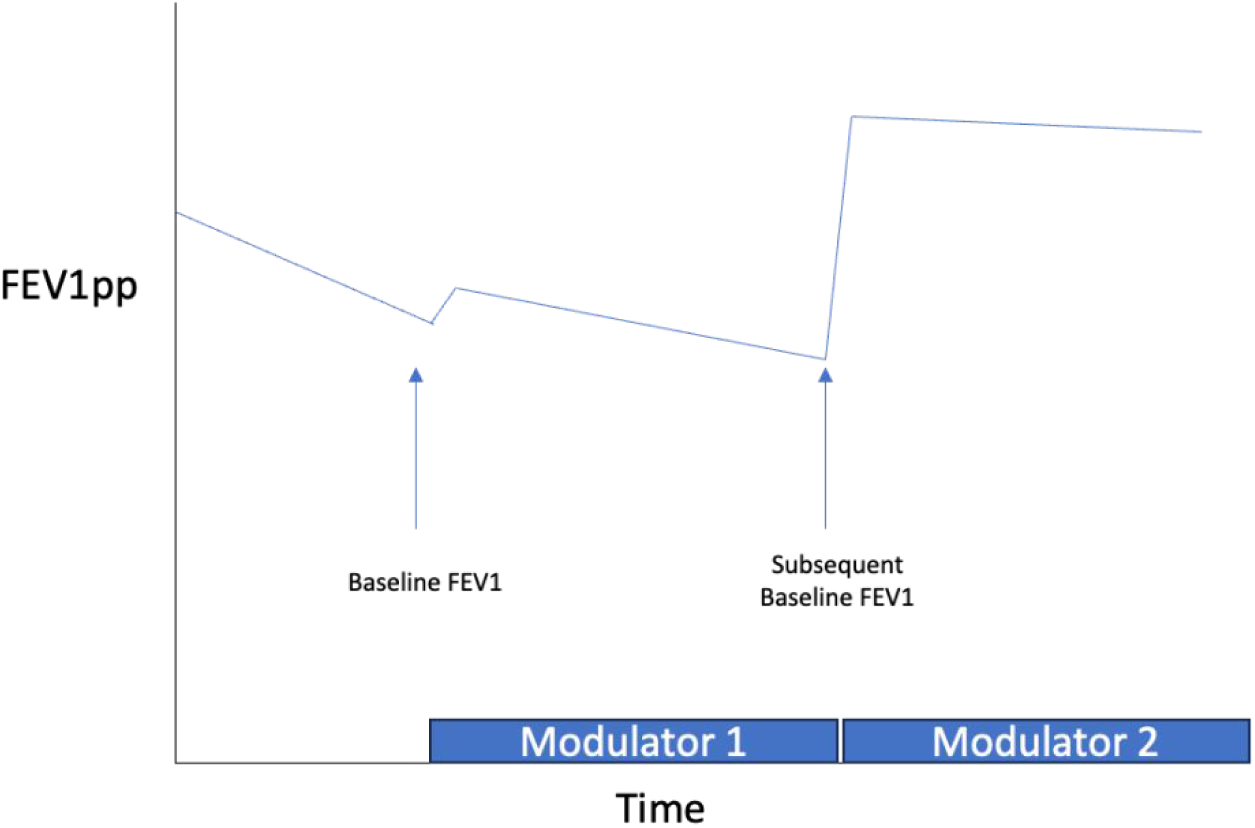
Schematic of baseline FEV1pp definition.

#### *In vivo* clinical response to modulator – sweat chloride (SC)

SC testing was performed by an accredited lab (NSW Health pathology) as part of routine clinical care of patients receiving a CFTR modulator at the SCH CF clinic. SC results were obtained from the participant’s medical record. Baseline SC was taken as the measurement prior to any CFTR modulator being taken by the patient. When a SC level immediately pre CFTR modulator treatment was not available, prior results from time of diagnosis were used (n = 2 teenage participants, SC results from infancy used as baseline). Post treatment SC results were obtained after at least 4 weeks of stable CFTR modulator dosing. Post treatment SC results for subsequent CFTR modulator treatments were compared to the baseline results, for the purpose of comparison to the participant’s *in vitro* data.

#### *CFTR* gene sequencing analysis and drug response relationship

The DNeasy® Blood & Tissue kit (Qiagen 69506) and the associated protocol was used to extract DNA from participant cells. Whole genome sequencing (WGS) were carried out at Ramaciotti Centre for Genomics (UNSW). DNA libraries were prepared using Illumina DNA prep kit and sequenced in 2×150bp paired-end format on Illumina iseq 100 system. Sequence data were processed using DRAGEN Germline Pipeline (v.4.1.7) for alignment, variant calling and quality control. Reads were mapped to alt-contig masked hg38 reference genome. Analysis was performed using the Gadi system of the National Computational Infrastructure (NCI). The CFTR gene was processed from promoter to the 3’ UTR terminus (GRCh38 coordinates chr7:117476071-117668665). VCF files of each participant for CFTR gene locus were merged using bcftools (V1.12)[6] excluding multiallelic records (-m none) and assuming reference homozygous genotypes (0/0) at invariable sites (-0). Variant consequence and clinical significance were annotated using Ensembl Variant Effect Predictor (V111) [7, 8]. VCF files were converted to GRCh37 coordinates using Ensembl Assembly Converter and phased using ShapeIT2 (v2, r904) [9] with the 1000 Genomes Phase 3 [10] genetic map and default parameters.

#### Pharmacogene profiling

To identify genetic variations influencing drug metabolism and transporter activity, WGS data was analyzed using Stargazer (Version 1.19.2)[11] to detect copy number variations (CNVs), single nucleotide polymorphisms (SNPs), and structural variants in pharmacogenes. Using patient BAM files as input, Stargazer’s –create-gdf-file function was employed to calculate read depth for all pharmacogenes, normalising against RYR1 as the control gene. After generating the GDF (GATK-DepthOfCoverage format) files, patient VCF files was used to call star alleles. The predicted activity levels of enzymes and transporters were categorized based on Stargazer output, focusing on four families; CYP (Cytochrome P450) enzymes, ABC (ATP-binding cassette) transporters, SLC and SLCO (solute carrier) families and UGT (UDP-glucuronosyltransferases). Activity levels were visualized in a heatmap in R using the pheatmap package (Version 1.0.12)[12] with color-coding to indicate variability in activity levels.

#### Creation of *in vitro* nasal epithelial cell cultures

Participants underwent brushing of the inferior nasal turbinates (Endoscan, 33009-SA McFarlane, Ringwood, VIC, Australia) to obtain epithelial cells as previously described [13, 14]. Briefly, collected cells were seeded in a collagen-I precoated flask (Advanced Biomatrix 5015) and co cultured with irradiated NIH/3T3 feeder cells. Human nasal epithelial cells (HNECs) were expanded using a conditional reprogramming culture (CRC) method, supplemented with ROCK (Rho kinase) inhibitor (10 μM) [15]. At 90% confluence, a double trypsin method was used to dissociate the HNECs, which were then cryopreserved at passage 1 in our CF biobank [13].

The cryopreserved HNECs were seeded (200-250,000/filter) onto collagen-I coated 6.5mm Transwell® membrane inserts (Sigma CLS3470) as previously described [13]. These HNECs were expanded using PneumaCult Ex Plus media (STEMCELL Technologies 05040) until confluent (4-6 days). The basal media was replaced with air liquid interface (ALI) differentiation media (STEMCELL Technologies 05001) and an ALI was created by removal of the apical media. The HNECs were maintained at ALI for 21 days with alternate basal media changes.

#### Cilia beat frequency measurement

Functional assessment of differentiated-HNEC cultures was conducted by measuring cilia beat frequency, as described previously [13]. Briefly, mature differentiated-HNEC cultures were imaged using high speed live cell imaging system (Eclipse Ti2-E, Nikon microscope, Andor Zyla 4.2 sCMOS camera, CFI S Plan Fluor ELWD 20×/0.45 objective) in an environmentally controlled chamber (37°C, 85% humidity and 5% CO2). Time-lapse images were acquired and CBF was analysed using a custom-built script in MATLAB (MathWorks, Natick, MA) [13, 16]. Six fields of view (512 x 512 pixels) were acquired from each transwell insert, with the mean of the six images being calculated as the result. A minimum of four filters were measured for each participant at baseline.

#### Quantification of *in Vitro* CFTR-mediated ion transport electrophysiology

CFTR-mediated ion transport was assessed by measurement of short circuit current (I_SC_) in the differentiated-HNEC cultures as described previously [15]. Briefly, differentiated-HNEC cultures were pre-incubated with CFTR correctors (3 µM LUM (VX-809; Selleckchem 1565 [17]), 5 µM TEZ (VX-661; Selleckchem S7059 [18], or a cocktail of 18 µM TEZ and 3 µM elexacaftor (E) (VX-445; Selleckchem S8851) [19] for 48h prior to assessment of ion transport electrophysiology. I_sc_ measurements were taken under voltage-clamp conditions using VCC MC8 Ussing chambers (Physiologic Instruments, San Diego, CA). Data recordings were acquired using Acquire and Analyze (version 2.3) software (Physiologic Instruments, San Diego, CA). Experiments were performed under a basal to apical Cl-concentration gradient, created using 10 mM HEPES buffered Ringer solutions. Basal solution contained (mM): 145 NaCl, 3.3 K_2_HPO_4_, 10 N-2-hydroxyethylpiperazine-N-2-ethane sulfonic acid (HEPES), 10 D-Glucose, 1.2 MgCl_2_, and 1.2 CaCl_2_. Apical solution contained (mM): 145 Na-Gluconate, 3.3 K_2_HPO_4_, 10 HEPES, 10 D-Glucose, 1.2 MgCl_2_, 1.2 CaCl_2_. Ringer solutions were continuously gassed with 95% O_2_-5% CO_2_ and maintained at 37°C. Following a 30 minute stabilisation period, differentiated-HNEC cultures were treated with pharmacological compounds (in order): 100 µM amiloride (Sigma A7410, apical) to inhibit epithelial sodium channel (ENaC)-mediated sodium (Na^+^) flux, vehicle control 0.01% DMSO or 10 µM IVA (VX-770; Selleckchem S1144; apical) to potentiate cAMP-activated currents, 10 µM forskolin (Sigma F6886, basal) to induce cAMP activation of CFTR, 30 µM CFTR_inh_-172 ( Selleckchem S7139, apical) to inhibit CFTR-specific currents and 100 µM ATP (Sigma A2383, apical) to activate calcium-activated chloride currents (**Figure 5A**).

#### Statistical analysis

Change in clinical measurements of FEV1pp and SC were evaluated using a paired t test. Waterfall plots were used to visualise the variations in clinical response between individuals. Participants were stratified into two groups for analysis based on their baseline FEV1pp; Group 1: FEV1pp 40 – 90 Group 2: FEV1pp < 40 or > 90, although no participants had an FEV1pp less than 40 at baseline. The relationship between clinical and *in vitro* functional response (ΔIsc) to the first CFTR modulator was evaluated using multiple regression analysis, which included age, gender and an interaction with baseline FEV1pp. Receiver operating characteristic (ROC) curves were generated for *in vitro* functional response (ΔIsc) that identified predefined significant clinical response. A significant increase in FEV1pp was considered an increased in absolute percent predicted greater than 5 percentage points. A significant improvement in SC was considered a decrease of greater than 20 mmol/L. To investigate the full data set available, which included responses to subsequent modulator treatment, we used generalised estimating equations with an unstructured covariance matrix to account for repeated measures. The model for change in FEV1pp included fixed effects for age, gender, CFTR modulator, baseline FEV1pp group, *in vitro* functional response (ΔIsc) and an interaction between baseline FEV1pp group and *in vitro* functional response (ΔIsc). The model for change in SC did not include baseline FEV1pp group. Residuals were checked, and sensitivity analysis run when model assumptions were not met by the data. Statistical analysis was performed with GraphPad Prism software (v9.5.1) and R (v4.2.2). A *p*-value of less than 0.05 was considered statistically significant.

## Notes

### Funding Statement

This work was supported in part by Sydney Childrens Hospitals Foundation and an Australian National Health and Medical Research Council grant (NHMRC_APP1188987), Rebecca L. Cooper Foundation project grant, Cystic Fibrosis Australia, The David Millar Giles Innovation Grant, and Luminesce Alliance Research grants. LF is supported by the Rotary Club of Sydney Cove-Sydney Childrens Hospitals Foundation and UNSW postgraduate award scholarships. KA is supported by an Australian Government Research Training Program Scholarship. HRP is supported by the Australian National Health and Medical Research Council grant (NHMRC_APP2021172). EKS-F is supported by the National Health and Medical Research Council (Grant ID: APP1157287), Cure4CF, Cystic Fibrosis Foundation (SCHNEI24I0) and The University of Melbourne. SAW is supported by UNSW Scientia program and the Australian National Health and Medical Research Council (NHMRC_APP1188987).

### Author Declarations

This study was approved by the Sydney childrens Hospital Network (SCHN) Ethics Review Board (HREC/16/SCHN/120). Written informed consent was obtained from all participating subjects and/or their legal guardians. Participants were recruited from the Sydney Childrens Hospital (SCH) cystic fibrosis (CF) clinic if they; 1) had an F508del homozygous CFTR genotype, 2) commenced treatment with a CFTR modulator with ongoing follow up by the CF team and 3) completed spirometry testing pre and post modulator treatment according to the American Thoracic Society (ATS) criteria. All necessary patient/participant consent (including consent to publish) has been obtained and the appropriate institutional forms have been archived and any patient/participant/sample identifiers included were not known to anyone (e.g., hospital staff, patients or participants themselves) outside the research group and cannot be used to identify individuals.

